# Clinical malaria incidence following an outbreak in Ecuador was predominantly associated with *Plasmodium falciparum* with recombinant variant antigen gene repertoires

**DOI:** 10.1101/2021.04.12.21255093

**Authors:** Shazia Ruybal-Pesántez, Fabian E. Sáenz, Samantha Deed, Erik K. Johnson, Daniel B. Larremore, Claudia A. Vera-Arias, Kathryn E. Tiedje, Karen P. Day

## Abstract

To better understand the factors underlying the continued incidence of clinical episodes of falciparum malaria in E-2020 countries targeting elimination, we have characterised *Plasmodium falciparum* disease transmission dynamics after a clonal outbreak on the northwest coast of Ecuador over a period of two years. We apply a novel, high-resolution genotyping method, the “*var*code” based on a single PCR to fingerprint the DBLα region of the 40-60 members of the variant surface antigen-encoding *var* multigene family. *Var* genes are highly polymorphic within and between genomes, with *var* repertoires rapidly evolving by outcrossing during the obligatory sexual phase of *P. falciparum* in the mosquito. The continued incidence of clinical malaria after the outbreak in Ecuador provided a unique opportunity to use *var*codes to document parasite microevolution and explore signatures of local disease transmission on the time scale of months to two years post-outbreak. We identified nine genetic *var*codes circulating locally with spatiotemporal parasite genetic relatedness networks revealing that diversification of the clonal outbreak parasites by sexual recombination was associated with increased incidence of clinical episodes of malaria. Whether this was due to chance, immune selection or sexual recombination per se is discussed. Comparative analyses to other South American parasite populations where *P. falciparum* transmission remains endemic elucidated the possible origins of Ecuadorian *var*codes. This analysis demonstrated that the majority of clinical cases were due to local transmission and not importation. Nonetheless, some of the *var*codes that were unrelated to the outbreak *var*code were found to be genetically related to other South American parasites. Our findings demonstrate the utility of the *var*code as a high-resolution surveillance tool to spatiotemporally track disease outbreaks using variant surface antigen genes and resolve signatures of recombination in an E-2020 setting nearing elimination.

## Introduction

In 2016 the WHO identified 21 countries, known as the E-2020 countries, having the potential to eliminate local transmission of malaria by 2020^1^, with seven of them in Latin America at the time (Paraguay and El Salvador have since been declared malaria-free). Malaria transmission in many of these E-2020 countries is epidemic/unstable with risks of outbreaks and resurgent malaria ^2–4^. The elimination target requires a local reduction to zero incidence of indigenous cases. As progress is made towards elimination in these countries, it becomes critical to understand risk factors for disease transmission while maximizing limited funds and resources to examine individual cases. Ecuador is one of the five remaining E-2020 countries in Latin America. National malaria elimination efforts have largely focused on tropical areas, specifically the northwest coast and the Amazon region. These areas border non E-2020 countries, Colombia and Peru, that still have endemic transmission. Thus, rather than low transmission, Ecuador has epidemic/unstable *P. falciparum* transmission with *P. vivax* the dominant species causing clinical malaria infections.

Clinical cases caused by *P. falciparum* in Ecuador are mostly concentrated in the northwest coast, where a steady decrease in annual reported clinical cases was observed from 2009 (161 cases) until late 2012 (44 cases)^5–7^. A localized outbreak from November 2012 until November 2013 in Esmeraldas City documented 151 *P. falciparum* cases, the majority of cases (76%) in the northwest coast and Amazon region during this time period^8^. Population genetic analyses using microsatellite markers demonstrated that this outbreak was caused by the clonal expansion (i.e., single-source) of a residual parasite lineage previously reported on the north coast of Peru and on the coast of Ecuador^8^. Immediately after the outbreak, 31 cases were reported in 2014 and 189 cases in 2015 ^5, 9,10^. It was unclear whether the clonal outbreak had contributed to *P. falciparum* clinical cases since 2013, with the factors underlying sustained disease transmission in Ecuador poorly understood. Moreover, Ecuador was categorized as “off-track” in the most recent E-2020 progress report and has not met the elimination target due to malaria resurgence^11, 12^. This points to an urgent need to better understand disease transmission patterns and whether clinical cases are due to imported or locally-acquired parasites.

Molecular surveillance of malaria in Latin America has focused on non E-2020 countries, e.g. Colombia, Brazil, Peru and Panama where malaria is endemic and transmission is moderate to low^8, 13–19^. This surveillance has relied on genotyping neutral molecular markers such as microsatellites or single-nucleotide polymorphisms. However, neutral markers, considered relatively slowly evolving, may not provide enough discriminatory resolution to define parasite evolution in relevant epidemiological time (1-2 years) in areas of low or epidemic and unstable transmission^20, 21^. Genes under selection can provide an alternative but complementary view of microevolution and they are key to examine population adaptation ^22^.

In this context, the microbiological paradigm for the surveillance of diverse pathogens is to study transmission dynamics using the genes encoding the major surface antigen to reflect pathogen transmission dynamics in relation to immune selection^23, 24^. The failure to observe this paradigm in the malaria field has stemmed from both the misconception that neutral markers track transmission dynamics in relation to immunity and the fact that the genetics of the *var* genes encoding the major surface antigen of the blood stages (PfEMP1) are considered “too complex”. Single copy antigen genes under much less selection have become an alternative^25^. Instead, we have embraced the complexity of the *var* system to understand the population genetics of *var* genes^23, 26–31^ in multiple locations in Africa, South America, and globally to present here a way to explore transmission dynamics through the lens of these immune evasion genes.

Each genomic *var* repertoire of *P. falciparum* consists of 40 to 60 members of the *var* multigene family with evidence that South American parasites may have fewer *var* genes, e.g. 42 *var* genes (excluding *var2csa)* reported in the Honduran laboratory strain HB3 after whole genome sequencing and assembly ^32, 33^. Our analysis of a South American dataset with a global dataset also showed fewer *var* genes in South American isolates as did a Brazilian data set ^31, 34^. As *var* genes lie on almost all of the 14 chromosomes of the haploid *P. falciparum* genome^35^ they undergo independent assortment during conventional meiosis to create new *var* repertoires as recombinants of parental *var* repertoires. During infection, differential switching between these genes in a repertoire enables immune evasion in the human host^36, 37^ thereby regulating parasite density and risk of clinical infection^38^. Indeed, several serologic studies examining anti-PfEMP1 antibody responses in naturally-infected children have shown that the acquisition of anti-PfEMP1 protective immunity is variant-specific and susceptibility to clinical episodes is dependent on gaps in the individual’s pre-existing antibody repertoire^38–47^.

A body of work defining the population genomics of the *var* multigene family by ourselves and others^48^ has demonstrated that *var* genes provide a promising biomarker for surveillance over epidemiological time scales (1-10 years). *Var* DBLα sequences can resolve population structure at global, continental, regional and local scales in areas of varying endemicity in Africa, Oceania, and South America^23, 26–31, 49^. Characterization of these major surface antigen genes also provides information about circulating repertoires of variants to enable measurement of immune responses. Translation of *var* population genetics in different epidemiological settings leads us to propose a new surveillance approach we call *var*coding. This is essentially a fingerprinting method to describe the diversity of these genes within and between human hosts as well as repertoire diversification by sexual recombination, with Bayesian statistics to account for missing data (Fig. 1).

**Figure 1.**
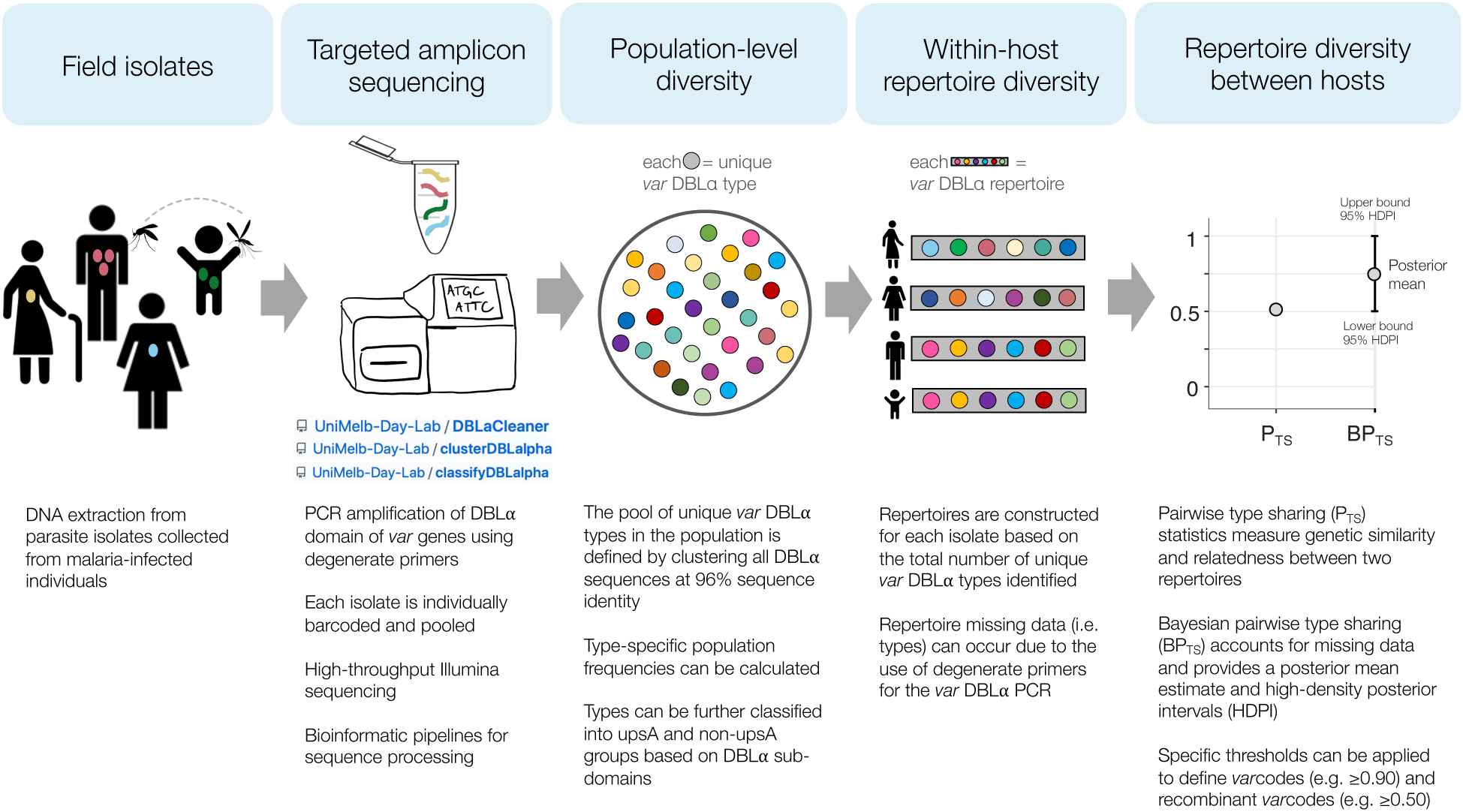
Schematic diagram of the *varcoding* approach. For more details about each step, see Methods.

Defining the 2012-2013 clinical disease outbreak in Ecuador as a baseline, we applied the *var*code methodology to characterize the transmission dynamics of clinical *P. falciparum* cases during and up to two years after the outbreak. Reconstruction of parasite genetic relatedness networks in space and time revealed that parasites with recombinant *var* repertoires were predominantly associated with the continued incidence of clinical cases after the outbreak. Further comparative analyses to published data from South American isolates^28^ elucidated possible origins of Ecuadorian parasites, demonstrating that the majority of clinical cases were due to local transmission and not importation. Our findings also demonstrate the translational application of the *var*code as a high-resolution tool to detect, spatiotemporally track immune evasion genes as well as resolve signatures of recombination in E-2020 or low transmission settings.

## Results

*Var*coding was completed for 58 *P. falciparum* isolates that were collected between 2013 and 2015 in endemic areas of the northwest coast and Amazon region of Ecuador (Fig. 2) from individuals of all ages presenting with malaria symptoms and confirmed *P. falciparum*-positive by microscopy or rapid diagnostic tests. These isolates represent 21% of the total cases reported in 2013, 61% in 2014 and 3% in 2015 (60% of the cases reported in January, 7% in May and 8% in November). For more details on the sampling scheme and bioinformatics workflow see Methods and Supplementary Fig. S1. Overall, we identified 195 unique *var* DBLα variants or types from 2,141 DBLα sequences from the 58 isolates, representative of the diversity circulating in Ecuadorian *P. falciparum* populations, as indicated by accumulation curves approaching a plateau (Fig. S2).

**Figure 2.**
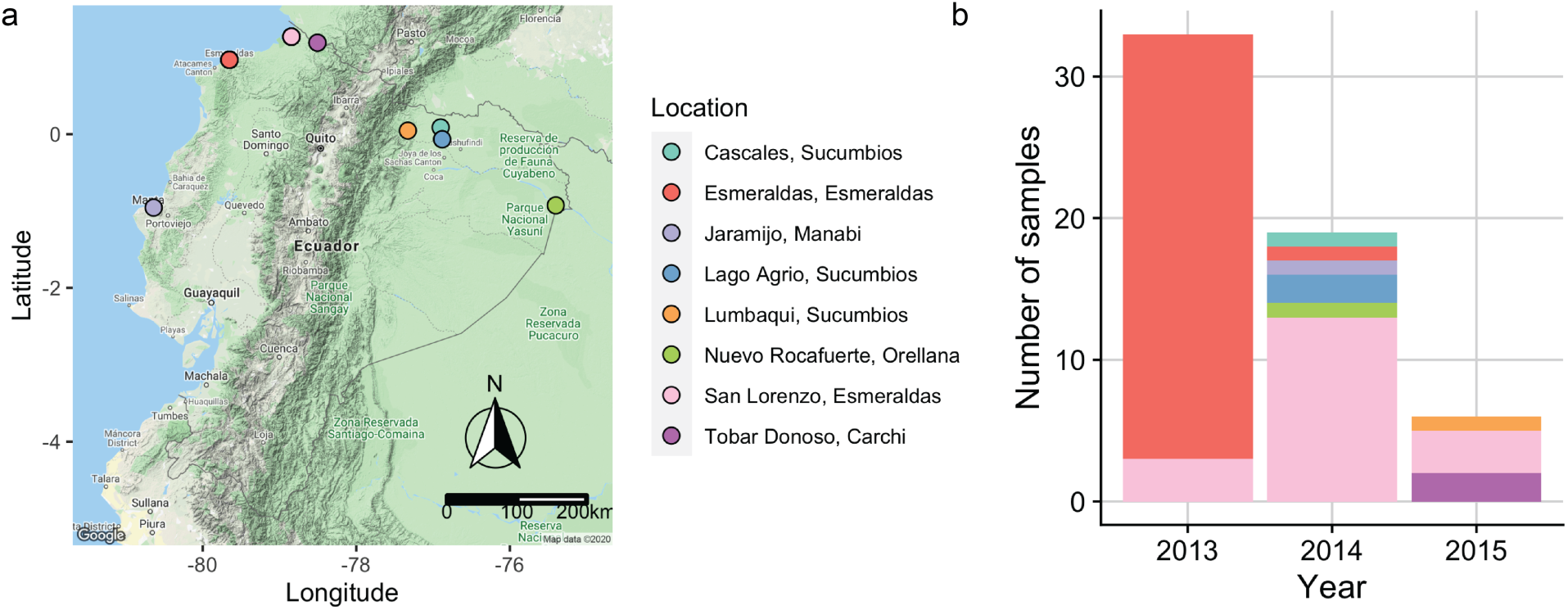
Sampling of *P. falciparum* isolates across endemic areas of Ecuador and over a period of three years (2013 – 2015). (a) Map of Ecuador depicting the sampling locations during the study. (b) Bar plot showing the number of *P. falciparum* positive samples collected in each year and their respective sampling locations. Every location is indicated with a different color.

### Defining *var*codes circulating in Ecuador

To define distinct *var*codes circulating locally in Ecuador, we estimated parasite genetic relatedness by calculating the similarity index, Pairwise Type Sharing (P_TS_) and a Bayesian version of the same (Bayesian pairwise type sharing; BP_TS_, see Methods). BP_TS_ estimates include statistical uncertainty due to differences in isolate repertoire size (i.e. the number of DBLα types identified in each isolate repertoire), enabling a rigorous examination of relatedness. The median repertoire size in Ecuadorian isolates was 37 DBLα types (range: 11-43 types, Table S1), which is in line with the expected number of *var* genes in the Honduran laboratory reference strain HB3 (n=42 *var* genes excluding *var2csa*, ^33^). Nonetheless, we used a uniform prior distribution for repertoire sizes between 40 and 50 types, which accounted for the possibility of missing data in total isolate repertoire sizes using degenerate primers for *var* DBLα PCR. For a further analysis to validate our PCR and sequencing methodology, including a further sub-classification of DBLα types into upsA and non-upsA groups and their respective isolate genomic proportions, see Methods and Supplementary Text 1.

We constructed genetic relatedness networks at a threshold of P_TS_ ≥ 0.90 to identify putatively identical genomes within the margin of error of detection of all DBLα types in an isolate. We confirmed this definition by comparing *var*codes derived from P_TS_ to those derived from posterior mean BP_TS_ estimates (Supplementary Fig. S3), and by examining the lower and upper bounds of the 95% highest density posterior intervals (HDPIs). This revealed nine genetically distinct *var*codes in this study with *var*code4, *var*code5, *var*code9 seen only once and at the other extreme 36 isolates had *var*code1 (Fig. 3a and Supplementary Fig. S4a). We were unable to confirm whether any of the Ecuadorian *var*codes were circulating pre-outbreak as samples were not available. As expected, the outbreak *var*code1 was clonal (indicated by HDPIs that included 1; Fig. S5d), as previously demonstrated by microsatellite genotyping^8^. It is worth noting that even though we only sampled 30 *P. falciparum* isolates in Esmeraldas City from 140 (21%) outbreak cases in 2013, the fact that the disease outbreak was clonal and represented 91% of the 154 total cases on the northwest coast and Amazon region during 2013, means that we characterized a representative sample of the cases from the outbreak and during that year.

**Figure 3.**
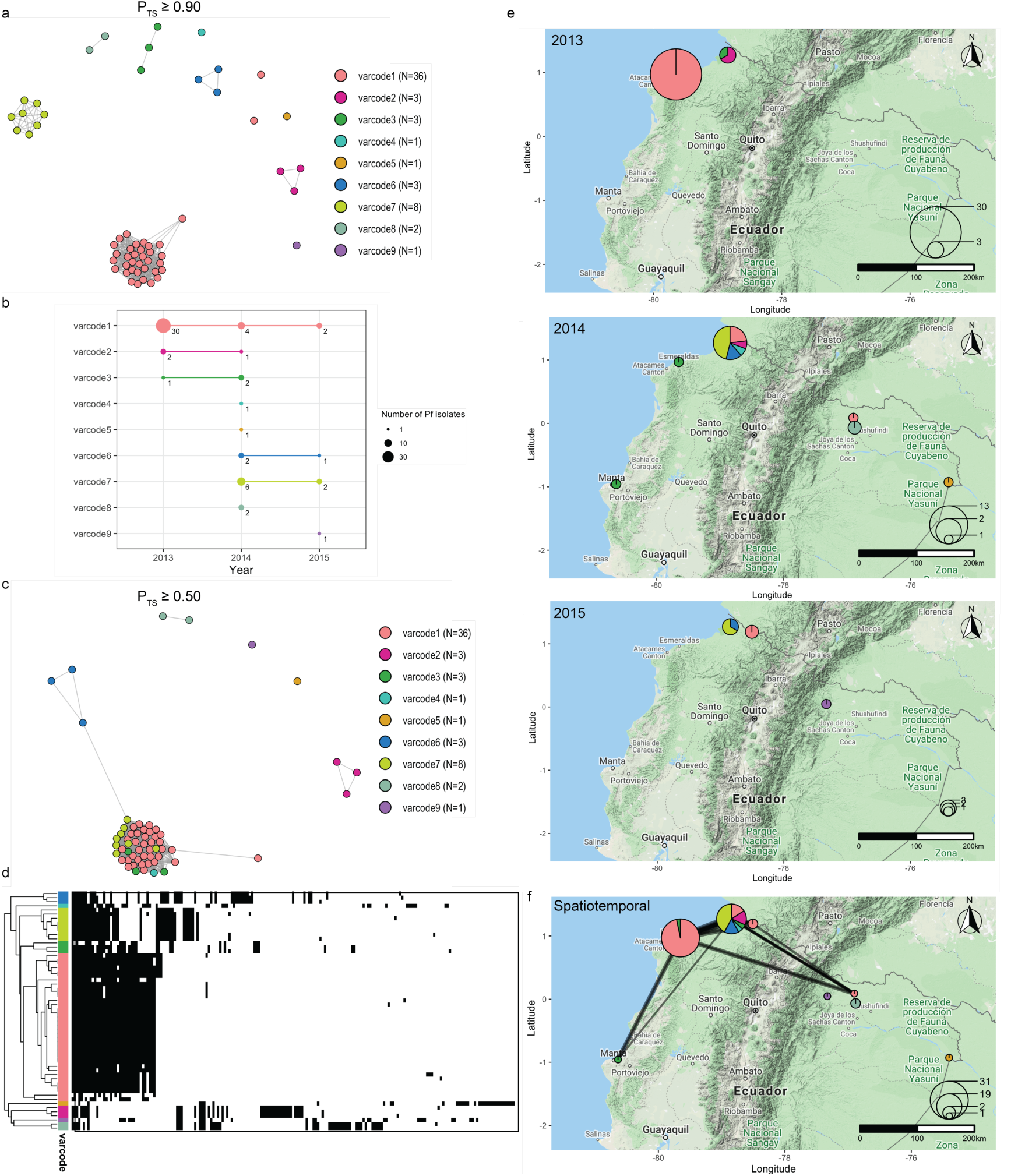
Reconstructing the disease transmission dynamics of *P. falciparum* reveals signatures of local transmission and sexual recombination after the outbreak. (a) A network visualization of the genetic relatedness of *P. falciparum* isolates at the threshold of PTS ≥0.90 to define *var*codes (see Methods). Every node represents a *P. falciparum* isolate and an edge represents the P_TS_ value between two particular nodes/isolates. Isolates that cluster together (i.e., connected by edges) are considered to have the same *var*code. Each color represents a different *var*code. Two *P. falciparum* isolates belonging to *var*code1 appear as outliers in the network due to undersampling of their DBLα types. N refers to the number of isolates. For comparison to BPTS estimates see Supplementary Fig. S3-5 and S8. (b) The number of *P. falciparum* isolates within each *var*code (i.e. size of circle) in each year and their persistence over time. The three *var*codes (*var*code1, *var*code2, *var*code3) identified in 2013 were identified again in 2014, and *var*code1 was identified again in 2015. Two *var*codes identified in 2014 (*var*code6, *var*code7) were also identified in 2015. (c) A network visualization of the genetic relatedness of *var*codes at the threshold of PTS ≥0.50 to discriminate between recombinant *var*codes and genetically distinct *var*codes. Isolates/*var*codes that cluster together (i.e., connected by edges) represent recombinants. (d) A clustered heatmap showing the genetic profiles of each *P. falciparum* isolate with rows representing each isolate and columns representing each DBLα type. Black and white denote the presence and absence of each type, respectively. Isolates that clustered together were more genetically similar (i.e., the same DBLα types were present). Similarly, *var*codes that clustered together were more genetically similar (e.g. recombinants). (e) The spatial distribution of *var*codes in each year, with the size of the circle representing the number of *P. falciparum* isolates sampled in a given location in each year and the pie chart depicting the proportion of each *var*code identified. (f) Spatiotemporal genetic relatedness network of *var*codes between 2013-2015. Every node represents a sampling location, the size of the circle represents the number of *P. falciparum* isolates sampled in each location, the pie chart depicts the proportion of each *var*code identified, and the weighted edges show genetically-related *var*codes (P_TS_ ≥0.50). (Online version: GIF animation depicts the spatiotemporal transmission dynamics chronologically during and after the outbreak).

For each pairwise comparison, we calculated the HDPI width as an estimate of uncertainty (analogous to calculating the width of a frequentist confidence interval) and found there was high confidence (HDPI width ≤0.2) for 94.6% (632/668) of pairwise BP_TS_ estimates (Supplementary Fig. S4b). Low confidence pairwise BP_TS_ estimates (HDPI width>0.2; max = 0.33) were found only between the two *var*code1 isolates with the smallest repertoire sizes (11 and 19 DBLα types; 36/36 comparisons). This lower statistical confidence follows naturally from under sampling of those isolates’ DBLα types. After aggregating the within-*var*code pairwise comparisons, there was high confidence in the definition of all *var*codes (Supplementary Fig. S5a-b).

For comparison of relatedness by microsatellite genotyping, we calculated Pairwise Allele Sharing (P_AS_) using previously published microsatellite data for the same isolates^8, 50^. Not surprisingly, 7 microsatellites did not resolve the nine *var*codes but only identified four, three of which were single parasite isolates, at the threshold of P_AS_ ≥ 0.90 (Supplementary Fig. S6a). Because two of the seven microsatellite markers were fixed in the population^8, 50^, we also generated the relatedness networks at the threshold of P_AS_ ≥ 0.80, which allowed for an error of detection of 1 microsatellite allele. However, the majority of isolates clustered at this threshold and only three *var*codes were resolved (Supplementary Fig. S6b). There were several pairwise comparisons (14%, 72/541 by P_TS_ and 6%, 31/541 by BP_TS_) where *P. falciparum* isolates were found to be identical by microsatellites (P_AS_ = 1) but much less related by *var* (P_TS_ or BP_TS_ ≤ 0.50, Supplementary Fig. S7). Nevertheless, the relationship between P_TS_ and P_AS_ scores was positively correlated (Pearson’s correlation coefficient = 0.757, *p* < 0.001, Supplementary Fig. S7a). Similarly, the relationship between BP_TS_ and P_AS_ estimates was also positively correlated (Pearson’s correlation coefficient = 0.779, *p* < 0.001, Supplementary Fig. S7b).

### Persistent association of clinical cases with the same *var*codes over time and large distances

We next explored whether parasites with the same *var*codes caused clinical episodes after the outbreak. Persistent disease transmission over time (Fig. 3b) and large distances (Fig. 3e-f) was found as the same *var*codes were identified post outbreak and sometimes in different sampling locations. The median time between first and last identification of the same *var*codes with any clinical case was 216 days or approximately 7 months (range = 190 – 823 days) during the study period. For example, the outbreak *var*code1 identified in Esmeraldas City was also identified in San Lorenzo, Esmeraldas (∼150km from Esmeraldas city) in 2013, then Cascales, Sucumbios (>300km away) in 2014, and then in Tobar Donoso, Carchi (∼150km away) in 2015 (Fig. 3b). This is in line with an earlier study using microsatellites that showed that *P. falciparum* isolates from the outbreak were genetically related to an Ecuadorian parasite isolate collected around 1990^8^. This result was also consistent with reports in Peru^51^ and Colombia^18, 52^ where identical parasites were shown to be circulating up to five and eight years after first identification, respectively.

### Spatiotemporal genetic relatedness networks reveal signatures of recombination

We next reconstructed spatiotemporal genetic relatedness networks to explore signatures of local disease transmission and parasite microevolution after the outbreak. We applied a P_TS_ threshold of ≥0.50 to detect recombinant and/or genetically-related *var*codes. By using the date and location where each isolate was collected, we animated the network chronologically to examine spatiotemporal disease transmission dynamics during the outbreak in 2013 and post-outbreak in 2014 and 2015 (Fig. 3c, 3f, GIF). This analysis resolved parasite microevolution relevant to disease transmission after the outbreak and revealed that four *var*codes (*var*code3, *var*code4, *var*code6, *var*code7) were recombinants of the outbreak *var*code1 (Fig. 3c). The remaining *var*codes did not cluster in the network and were genetically distinct. These patterns were confirmed using BP_TS_ estimates based on posterior means (Supplementary Fig. S8a) and their corresponding 95% HDPIs (Supplementary Fig. S5c-f and S8b-c). For the *var*codes that were recombinants and/or genetically-related (i.e., cluster in the networks in Fig. 3c and Supplementary Fig. S8a), 70.9% (433/611) of the HDPIs were confidently above the threshold of BP_TS_ ≥0.50 (Supplementary Fig. S5d). For *var*codes that were genetically-distinct (i.e., do not cluster in the networks), 98.4% (368/374) of the HDPIs were confidently below the threshold of BP_TS_ ≤0.50 (Supplementary Fig. S5f). As expected, the relatedness networks based on the 7 microsatellites did not resolve these patterns since all isolates were connected at the threshold of P_AS_ ≥ 0.50 and genetic relatedness was overestimated (Supplementary Fig. S9a). Indeed, we were not always able to discriminate recombinants from genetically distinct parasites since several pairwise comparisons (25%, 307/1232 by P_TS_ and 16%, 196/1232 by BP_TS_) that resulted in P_AS_ ≥ 0.50 were found to be less related by *var* (P_TS_ or BP_TS_ ≤ 0.50) (Supplementary Fig. S7). Furthermore, the majority of recombinants were classified as essentially clonal since they clustered together at the threshold of P_AS_ ≥ 0.80 (allowing for differences in one microsatellite allele, Supplementary Fig. S6b).

We were able to detect that *var*code3 was in fact a recombinant of the outbreak *var*code1, even though it was sampled in San Lorenzo during the same time period as the ongoing outbreak in 2013 (Fig. 3c, 3e). With the available data we were unable to confirm whether *var*code3 was ‘imported’ to San Lorenzo from Esmeraldas City. The most plausible explanation is that the recombination event occurred in San Lorenzo during the outbreak. Parasites with recombinant *var*code3 caused clinical episodes in multiple locations in 2014 (Fig. 3e). The other recombinant *var*codes (*var*code4, *var*code6, *var*code7) were identified after the outbreak in 2014 or 2015. Whether the recombinant *var*codes resulted from sexual recombination events between outbreak *var*code1 and genetically distinct parasites that were already circulating at low levels and/or in asymptomatic reservoirs in Ecuador (e.g. ^5^) or those that were previously imported could not be ascertained from the current study population. Interestingly, all the recombinant *var*codes were identified in San Lorenzo indicating this area is a transmission hot spot and a reservoir of parasites with diverse *var* repertoires (Fig. 3e).

To further visualize the genetic profiles of *var*codes and confirm the observed recombination signatures, we constructed a clustered heatmap based on the presence and absence of the 195 DBLα types across all isolates, such that isolates with similar genetic profiles cluster together (Fig. 3d). This analysis confirmed that in the case of isolates identified as recombinants of the outbreak *var*code1 (i.e., *var*code3, *var*code4, *var*code6, *var*code7), a proportion of outbreak DBLα types as well as different DBLα types were present. This is consistent with the generation of a new *var*code through outcrossing between two genomes with different *var* repertoires by conventional meiosis (i.e., approximately ≥50% sharing of DBLα types). By contrast, the genetic profiles of *var*codes 2, 5, 8, and 9 had different DBLα types, leading to the definition as parasites with genetically distinct *var*codes. When using 7 microsatellites, these recombination signatures in the genetic profiles were not identified (Supplementary Fig. S9b).

### Parasites with recombinant *var*codes most frequently caused *P. falciparum* clinical episodes following the 2012-13 outbreak

Prior to the outbreak there had been a steady decline in clinical cases, however, increased incidence of disease occurred after the outbreak. Therefore, we analyzed trends in the epidemiology of *P. falciparum* cases that occurred post-outbreak to understand if there were any key risk factors. Of the 25 individuals in our study that had a clinical *P. falciparum* episode after the outbreak in 2014 or 2015, we had age data for 18 individuals (72%, Table 1). There was no significant difference (Kruskal-Wallis test, *p* = 0.65) in the median age of individuals experiencing clinical episodes caused by parasites with the outbreak *var*code1 (19 years, range = 19 – 57 years, N = 3 patients), a recombinant of *var*code1 (i.e., *var*codes 3, 4, 6, or 7) (25 years, range = 17 – 58 years, N = 13 patients), or by a different *var*code (34.5 years, range = 32 – 37 years, N = 2 patients). A greater diversity of *var*codes (1-9 inclusive) was associated with incidence of clinical malaria post-outbreak than during the 2013 outbreak (*var*codes 1,2,3). Indeed in 2014, 79% of the cases sampled were caused by either parasites with the outbreak *var*code1 (21%) or with any of the four recombinants of *var*code1 (58%). The trend was similar in 2015 with 83% of the cases sampled caused by either parasites with the outbreak *var*code1 (33%) or with any of the four recombinants of *var*code1 (50%), although our sampling of reported cases in 2015 after January was limited. Importantly, overall we found that 80% of the cases we sampled after the outbreak were caused by either parasites with the outbreak *var*code1 (24%) or with any of the four recombinants of *var*code1 (56%), especially with varcode7. Thus, disease transmission after the outbreak in 2014 and into 2015 was predominantly associated with parasites with recombinant *var* repertoires of the outbreak clone.

**Table 1.** Epidemiological characteristics of study participants.

| Case | Month collected |  | Age range, years | Sex | Sampling location (County, Province) | Self-reported infection location* | varcode | Putative origin** |
| --- | --- | --- | --- | --- | --- | --- | --- | --- |
| EC1 | Jan 2013 | Outbreak | NA | NA | Esmeraldas, Esmeraldas |  | varcode1 | Local |
| EC2 | Jan 2013 | Outbreak | NA | NA | Esmeraldas, Esmeraldas |  | varcode1 | Local |
| EC3 | May 2013 | Outbreak | >30-40 | M | Esmeraldas, Esmeraldas |  | varcode1 | Local |
| EC4 | May 2013 | Outbreak | >30-40 | M | Esmeraldas, Esmeraldas |  | varcode1 | Local |
| EC5 | May 2013 | Outbreak | >50-60 | F | Esmeraldas, Esmeraldas |  | varcode1 | Local |
| EC6 | May 2013 | Outbreak | >30-40 | M | Esmeraldas, Esmeraldas |  | varcode1 | Local |
| EC7 | May 2013 | Outbreak | >50-60 | F | Esmeraldas, Esmeraldas |  | varcode1 | Local |
| EC8 | May 2013 | Outbreak | >10-20 | F | Esmeraldas, Esmeraldas |  | varcode1 | Local |
| EC9 | May 2013 | Outbreak | >10-20 | F | Esmeraldas, Esmeraldas |  | varcode1 | Local |
| EC10 | May 2013 | Outbreak | >10-20 | M | Esmeraldas, Esmeraldas |  | varcode1 | Local |
| EC11 | May 2013 | Outbreak | >20-30 | F | Esmeraldas, Esmeraldas |  | varcode1 | Local |
| EC12 | Jun 2013 | Outbreak | >10-20 | F | Esmeraldas, Esmeraldas |  | varcode1 | Local |
| EC13 | Jun 2013 | Outbreak | >10-20 | M | Esmeraldas, Esmeraldas |  | varcode1 | Local |
| EC14 | Jun 2013 | Outbreak | >40-50 | M | Esmeraldas, Esmeraldas |  | varcode1 | Local |
| EC15 | Jun 2013 | Outbreak | >40-50 | M | Esmeraldas, Esmeraldas |  | varcode1 | Local |
| EC16 | Jun 2013 | Outbreak | >10-20 | F | Esmeraldas, Esmeraldas |  | varcode1 | Local |
| EC17 | Jun 2013 | Outbreak | >10-20 | F | Esmeraldas, Esmeraldas |  | varcode1 | Local |
| EC18 | Jun 2013 | Outbreak | >20-30 | M | Esmeraldas, Esmeraldas |  | varcode1 | Local |
| EC19 | Jun 2013 | Outbreak | >40-50 | M | Esmeraldas, Esmeraldas |  | varcode1 | Local |
| EC20 | Jul 2013 |  | >20-30 | M | San Lorenzo, Esmeraldas |  | varcode2 | Local |
| EC21 | Jul 2013 |  | >10-20 | F | San Lorenzo, Esmeraldas |  | varcode2 | Local |
| EC22 | Jul 2013 | Outbreak | >10-20 | M | Esmeraldas, Esmeraldas |  | varcode1 | Local |
| EC23 | Jul 2013 | Outbreak | >0-10 | M | Esmeraldas, Esmeraldas |  | varcode1 | Local |
| EC24 | Jul 2013 | Outbreak | NA | F | Esmeraldas, Esmeraldas |  | varcode1 | Local |
| EC25 | Sep 2013 | Outbreak | >30-40 | M | Esmeraldas, Esmeraldas |  | varcode1 | Local |
| EC26 | Oct 2013 | Outbreak | >20-30 | F | Esmeraldas, Esmeraldas |  | varcode1 | Local |
| EC27 | Oct 2013 |  | >10-20 | F | San Lorenzo, Esmeraldas |  | varcode3 | Local |
| EC28 | Oct 2013 | Outbreak | >20-30 | M | Esmeraldas, Esmeraldas |  | varcode1 | Local |
| EC29 | Nov 2013 | Outbreak | >10-20 | M | Esmeraldas, Esmeraldas |  | varcode1 | Local |
| EC30 | Nov 2013 | Outbreak | >10-20 | M | Esmeraldas, Esmeraldas |  | varcode1 | Local |
| EC31 | Nov 2013 | Outbreak | >10-20 | F | Esmeraldas, Esmeraldas |  | varcode1 | Local |
| EC32 | Nov 2013 | Outbreak | >30-40 | M | Esmeraldas, Esmeraldas |  | varcode1 | Local |
| EC33 | Nov 2013 | Outbreak | >30-40 | M | Esmeraldas, Esmeraldas |  | varcode1 | Local |
| EC34 | Jan 2014 |  | NA | NA | Jaramijo, Manabi |  | varcode3 | Local |
| EC35 | Jan 2014 |  | >50-60 | F | San Lorenzo, Esmeraldas |  | varcode1 | Local |
| EC36 | Jan 2014 |  | >10-20 | M | San Lorenzo, Esmeraldas |  | varcode1 | Local |
| EC37 | Jan 2014 |  | >30-40 | M | San Lorenzo, Esmeraldas |  | varcode2 | Local |
| EC38 | Jan 2014 |  | >10-20 | M | San Lorenzo, Esmeraldas |  | varcode4 | Local |
| EC39 | Jan 2014 |  | >10-20 | M | San Lorenzo, Esmeraldas |  | varcode1 | Local |
| EC40 | Jan 2014 |  | >30-40 | F | Aguarico, Orellana | Peru | varcode5 | French Guiana/Peru/Venezuela |
| EC41 | Mar 2014 |  | >20-30 | M | San Lorenzo, Esmeraldas |  | varcode6 | Local |
| EC42 | May 2014 |  | >50-60 | F | Esmeraldas, Esmeraldas | Colombia | varcode3 | Local |
| EC43 | May 2014 |  | NA | NA | Cascales, Sucumbios | San Lorenzo, Esmeraldas | varcode1 | Local |
| EC44 | Jul 2014 |  | >30-40 | F | San Lorenzo, Esmeraldas |  | varcode7 | Local |
| EC45 | Aug 2014 |  | >20-30 | F | San Lorenzo, Esmeraldas |  | varcode7 | Local |
| EC46 | Aug 2014 |  | >20-30 | M | San Lorenzo, Esmeraldas |  | varcode7 | Local |
| EC47 | Aug 2014 |  | >50-60 | M | San Lorenzo, Esmeraldas |  | varcode7 | Local |
| EC48 | Aug 2014 |  | >20-30 | M | San Lorenzo, Esmeraldas |  | varcode7 | Local |
| EC49 | Sep 2014 |  | NA | F | Lago Agrio, Sucumbios | Colombia | varcode8 | Local |
| EC50 | Oct 2014 |  | NA | NA | Lago Agrio, Sucumbios | Colombia | varcode8 | Local |
| EC51 | Oct 2014 |  | >30-40 | M | San Lorenzo, Esmeraldas | Colombia | varcode6 | Local |
| EC52 | Dec 2014 |  | >20-30 | M | San Lorenzo, Esmeraldas |  | varcode7 | Local |
| EC53 | Jan 2015 |  | NA | NA | Lumbaqui, Sucumbios | Colombia | varcode9 | Colombia |
| EC54 | Jan 2015 |  | >20-30 | M | San Lorenzo, Esmeraldas |  | varcode7 | Local |
| EC55 | Jan 2015 |  | >10-20 | M | San Lorenzo, Esmeraldas |  | varcode7 | Local |
| EC56 | Nov 2015 |  | >10-20 | F | San Lorenzo, Esmeraldas |  | varcode6 | Local |
| EC57 | May 2015 |  | NA | NA | Tobar Donoso, Carchi |  | varcode1 | Local |
| EC58 | May 2015 |  | NA | NA | Tobar Donoso, Carchi |  | varcode1 | Local |

NA refers to no data collected for that particular epidemiological variable.
\*The self-reported infection location was recorded at the time of sample collection based on the answers provided by the study participants.
\*\*The putative origin as determined by varcoding.

### Comparative analyses to South America: elucidating possible origins of Ecuadorian *var*codes

We next examined the possible origins of the *var*codes circulating locally in Ecuador by comparing to published data^28^ from 128 *P. falciparum* isolates collected from neighboring South American *P. falciparum* populations (Fig. 4a). When we combined the sequences from Ecuadorian isolates with those previously sequenced, we identified 543 unique *var* DBLα types in South America (compared to 458 in ^28^). This is largely representative of the *var* diversity circulating in South American *P. falciparum* populations, as indicated by sampling accumulation curves approaching saturation (Supplementary Fig. S10). *Var*coding resolved a total of 97 *var*codes in South America (P_TS_ ≥ 0.90), with those identified in each country ranging from 9 in Ecuador and Venezuela, to 56 *var*codes in French Guiana (Supplementary Fig. S11). The number of *var* DBLα types per isolate repertoire was low in all countries and indicative of only one *P. falciparum* genome infecting an individual (i.e. repertoire size ≤ 60, Fig. 4b, Supplementary Table S1). We found that the median number of *var* DBLα types in each isolate ranged from 36.5 in Venezuela to 48.0 in French Guiana and the maximum number of *var* DBLα types per isolate was 42 or 43 in all countries except French Guiana that appeared to have more multi-genome infections (max = 92). The median repertoire size was in the range of the number of *var* genes seen in the genome of the Honduran laboratory reference strain HB3 (N=42 excluding *var2csa*, ^32, 33^). An unrooted phylogenetic neighbor-joining tree revealed distinct clusters of genetically-related isolates in South America generally clustering by country (Fig. 4c), consistent with previous analyses in the region demonstrating geographic population structure^28^.

**Figure 4.**
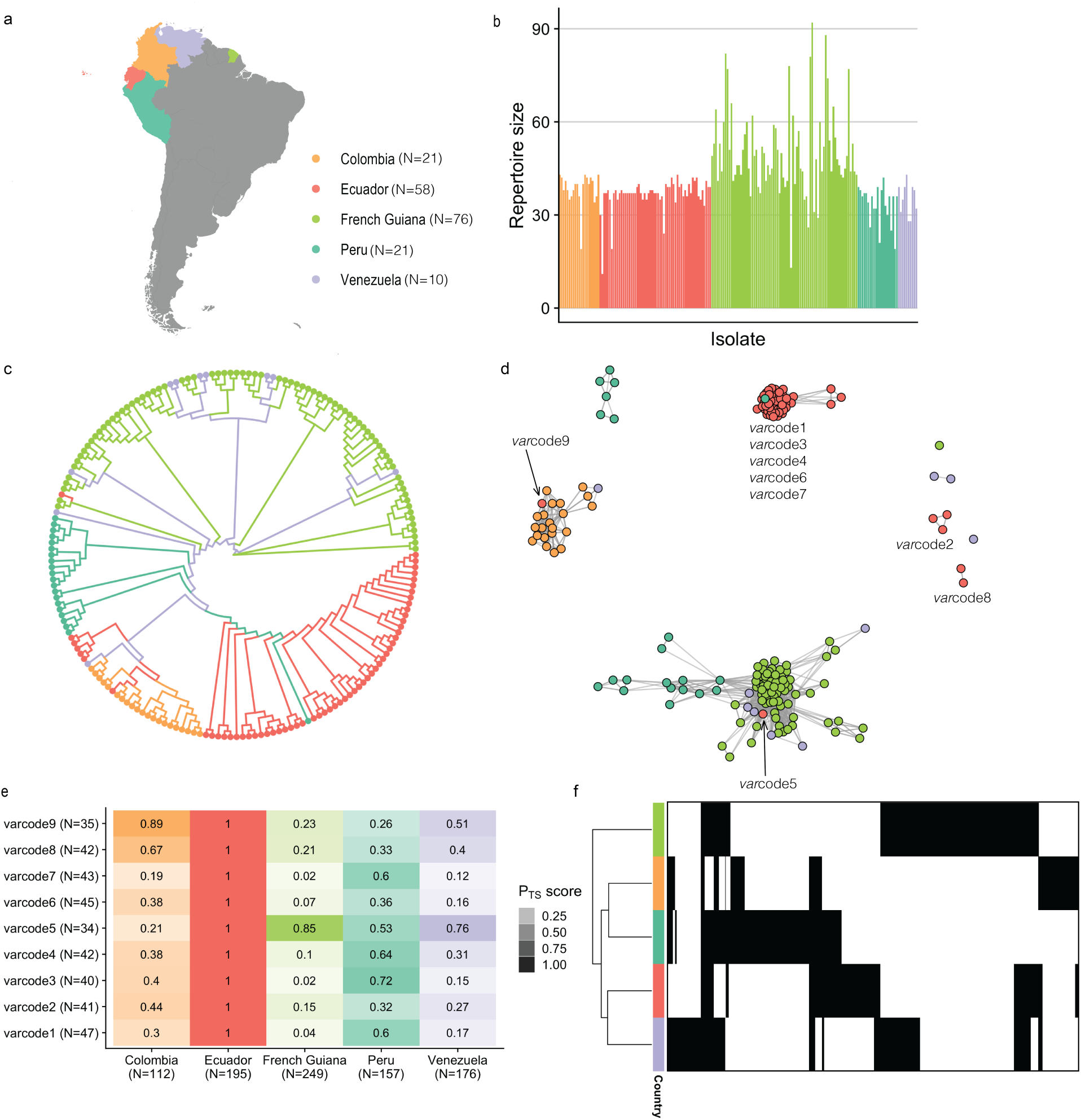
Relatedness networks in South America provide insights into the origins of the Ecuadorian *var*codes and diversity patterns in South America. (a) A map showing the study sites across South America. The dates of sample collection in these countries occurred from 2002 to 2008, around five to thirteen years prior to the sample collection of the Ecuadorian isolates. N refers to the number of isolates. (b) Repertoire sizes are shown for each *P. falciparum* isolate, from which multiplicity of infection can be inferred (i.e., ∼40-60 *var* DBLα types per *P. falciparum* genome). (c) An unrooted neigh14bor-joining tree shows the relatedness patterns among South American *P. falciparum* isolates. (d) A network visualization of the genetic relatedness of South American *var*codes at the threshold of PTS ≥0.50. Due to the differences in time points and sample collections, for this analysis a genetically-related parasite did not necessarily reflect a recent transmission event. Every node represents a *P. falciparum* isolate and an edge represents the PTS value between two particular nodes/isolates. Isolates/*var*codes that cluster together (i.e., connected by edges) are genetically-related. (e) Relatedness of Ecuadorian *var*codes to *P. falciparum* populations from South America. The relatedness of each *var*code was measured by first concatenating all possible *var* DBLα types that were identified in the *P. falciparum* isolates comprising each *var*code as well as concatenating all *var* DBLα types that were identified in each country. Then PTS was calculated between each *var*code and each country. The color gradient denotes the PTS value for a particular comparison, with darker shades showing higher relatedness and the different colors corresponding to the different country comparisons. N refers to the number of *var* DBLα types. (f) A clustered heatmap showing the patterns of diversity across South America, with a row representing all the types identified in a particular country, and columns representing each of the 543 DBLα types. Black and white denote the presence and absence of each type, respectively. The total number of unique *var* DBLα types identified in each country ranged from 112 in Colombia to 249 in French Guiana. Countries that cluster together share more *var* DBLα types.

To examine whether any of the parasites with Ecuadorian *var*codes were genetically related to other South American parasites, we constructed regional spatiotemporal relatedness networks and applied our threshold of P_TS_ ≥ 0.50 (Fig. 4d). We identified a genetically-related Peruvian *P. falciparum* isolate that clustered with the outbreak and recombinant *var*codes (P_TS_ = 0.66-0.75 with *var*code1 isolates). This is consistent with previous analyses using microsatellites showing the outbreak source was possibly a residual parasite lineage circulating in Peru in 1999-2000 and in Ecuador in the early 1990s^8, 13^. The fact that these isolates did not cluster with anything else suggests the outbreak may have been caused locally due to circulating parasites in unidentified reservoirs. *Var*code5, which was sampled in the Amazon had a very different genetic profile to the other *var*codes (≥83% unique types, Fig. 3d). It is worth noting that the parasite isolate with *var*code5 also had a different genetic background with respect to its microsatellite haplotype as compared to the rest of the Ecuadorian isolates (3 of 7 unique microsatellite alleles). The epidemiological data for the putative infection location was recorded as possibly imported from Peru (EC40, Table 1). However, our analysis would suggest that this *var*code is more related to *P. falciparum* populations from French Guiana and Venezuela. Nonetheless, Peruvian isolates also clustered with *P. falciparum* isolates from French Guiana even though they did not cluster with *var*code5 and we found that 53% of the types identified in *var*code5 were also identified in Peruvian parasites (Fig. 4e). We were able to determine a putative importation of a Colombian parasite (*var*code9, EC53, Table 1, Fig. 4d). The *var*codes 2 and 8 did not cluster with any other South American isolate suggesting parasites with these *var*codes may be local to Ecuador. The epidemiological data for the putative infection location for *var*code8 was recorded as Colombia (EC49 and EC50, Table 1). Although it did not cluster with Colombian *P. falciparum* isolates in our network analysis, we found that 67% of the types identified in *var*code8 (collapsing all 3 isolates) were also identified in Colombian parasites (Fig. 4e). For *var*code2, we found that 44% of the types identified in *var*code2 were also identified in Colombia (Fig. 4e), providing evidence that parasites with *var*code2 and *var*code8 may represent residual parasites previously or recently imported from Colombia.

### Conservation of individual DBLα types in space and time

Of significance, we found the same *var* DBLα types (56% of types identified in Ecuador, N=110/195, Fig. 4f) and *var*codes that were genetically related to other South American parasites and sampled up to 13 years before (Fig. 4d-e) were contributing to local transmission in Ecuador. Indeed, conservation of the DBLα types in all *var*codes was observed in space and time to varying degrees (Fig. 4e). We also found conservation of DBLα types on a continental scale with the number of the same types identified in any two or three countries being 124 (23%) and 59 (11%), respectively (Fig. 4f). Moreover, even DBLα types that were identified in relatively low frequencies in Ecuador (i.e., identified in a few *P. falciparum* isolates) were also identified in the South America dataset, often at relatively high frequencies in other countries (Supplementary Fig. S12). These patterns are consistent with some parasite *var*codes being putative importations and/or highly-related to parasites in other countries, as described previously.

### No evidence of recombination in any DBLα type over the course of the outbreak

The fact that the outbreak *var*code1 was clonal provided a unique opportunity to examine the conservation of its 47 individual DBLα types after transmission to multiple human hosts over the course of the outbreak and up to two years after (N=36 infections). DBLα types are very variable, with the average pairwise identity of the sequences encoding this domain being approximately 42%^33^. The cut-off of 96% sequence identity in our pipeline has been routinely used to identify the same DBLα types within the limits of sequencing errors and PCR artifacts (^27, 53^, Methods). Our data show that the same DBLα types were conserved in isolates with *var*code1 over the course of the outbreak (N=30 infections) and up to two years after the outbreak in the other six cases with *var*code1 (Supplementary Fig. S13a). In total, we report 1,283 independent observations of the same 41 DBLα types identified in 36 cases with *var*code1 assessed over 2 to 3 years (any type was identified in a median of 34 infections, ranging from 1 to all 36 infections) (Supplementary Fig. S13a). Given our threshold of P_TS_ ≥0.90 to identify putatively identical *var* DBLα repertoires, only 10 of the 47 DBLα types found in the outbreak clone were not found in the majority of the isolates with *var*code1 (median frequency = 1, max = 6, Supplementary Fig. S13a). Four of these were identified in the cases with recombinant *var*codes (median frequency = 17.5, range = 6-19), two of which were also identified in the Peruvian isolate that clustered with the outbreak/recombinants in Fig. 4d, suggesting they are also conserved. The other 6/10 were singletons in the entire dataset (including the South America data) and could be a consequence of the PCR with degenerate primers and/or low-quality DNA.

Among the 15 cases with recombinant *var*codes (*var*codes 3,4,6, and 7), we report 339 independent observations of 39 of the 47 outbreak DBLα types. There were 26-27 outbreak types shared between *var*code1 and the recombinant *var*codes 3,4, and 7, and 17 types between *var*code1 and *var*code6 (Supplementary Fig. S13b). These findings highlight the conservation of DBLα types of the outbreak clone in space and time, a pattern further supported by the fact that 31 of 47 outbreak DBLα types were found among previously recorded South American isolates (in 2002 to 2008^28^; see Supplementary Fig. S13c). This evidence points to conservation of these types despite the many transmission events that must have occurred between 2002 and 2013-15 across South America. We found that the highest number of outbreak DBLα types were identified in Peru (N=28, 60%) followed by Colombia (N=14, 30%) and Venezuela (N=8, 17%), and the lowest in French Guiana (N= 2, 4%, Supplementary Fig. S9c).

## Discussion

Our investigation of the spatiotemporal incidence of clinical episodes of *P. falciparum* in Ecuador by *var*coding supports the view that disease transmission after the 2012-2013 outbreak was sustained predominantly by Ecuadorian parasites. We observed persistence of the outbreak clonal lineage (identified with the same *var*codes) and outcrossing of this lineage with other locally circulating parasites causing clinical cases, rather than recent importation of clonally transmitted parasites. Our results point to the need for resources to be focused locally in Ecuador to uncover the circulating asymptomatic reservoirs of infection identified by *var*code recombination events. A role for human mobility must also be considered in the spread of *P. falciparum* as parasites with the same *var*codes were also observed across large distances (∼150-300km).

San Lorenzo was found to be a transmission hotspot likely due to mining activities and occupation-related travel in these areas, as well as its proximity to the Ecuador-Colombia border^3, 50, 54–56^. These findings point to San Lorenzo for both genetic surveillance and targeted interventions. Importantly, our results provide strong evidence for ongoing transmission in Ecuador and provide the first baseline characterization of *P. falciparum* antigenic diversity and parasite *var*codes circulating in the northwest coast and Amazon regions of Ecuador. It is worth noting that several recent outbreaks have occurred in the same areas of Ecuador (in 2016^10^, 2018^57^, 2019^6^ and 2020^58^) with malaria transmission remaining unstable rather than endemic.

We demonstrate that 58% of clinical cases sampled in 2014 immediately after the outbreak in Ecuador were caused by parasites with recombinant *var*codes. The same trend was observed in 2015, although our sampling was limited relative to the number of reported cases for the months sampled. We ask is this a chance finding or a consequence of immune evasion? We hypothesize that these parasites with recombinant *var*codes have new combinations of *var* DBLα types that may provide an immunological advantage in a population with variable levels of immunity, as would be expected during elimination campaigns^59^.

Network analyses and computational models combining evolution and epidemiology point to variant-specific immune selection defining *var* population structure^60, 61^. In scenarios where transmission is low and there is limited *var* diversity, the chance of recombination is also relatively low and thus immune selection is weaker. Selection in the context of an E-2020 country like Ecuador is likely variable due to the recent transition from moderate to epidemic transmission over the last 15 years^1^ with declining population immunity as a consequence of approaching elimination. Low population immunity would select for circulating clones leading to high similarity between *var* repertoires. Here, the signatures of recently recombined genomes with the outbreak clone that we detect in four *var*codes were clearly associated with the continued incidence of clinical disease after the outbreak. Thus parasites with recombinant *var*codes may present a risk factor for disease. We hypothesize that they may be better able to evade variant-specific immunity to PfEMP1 variants expressed by the outbreak clone by sharing 50% or less of the DBLα types of the outbreak clone. The availability of *var* DBLa sequences provides the potential to test this hypothesis by serological methods that detect variant-specific immunity. In this context, inferences based on neutral markers do not give information about immune evasion genes and variant-specific immunity, whereas *var*codes identify circulating variants to which individuals can become immune. e.g. as was done in serologic studies in Papua New Guinea^62^ and the Brazilian Amazon^63^.

A previous study that included some of the same *P. falciparum* isolates from Esmeraldas City and San Lorenzo in the northwest coast described three main genetic clusters based on microsatellite genotyping^50^. In contrast, when considering the same isolates, *varcoding* resolved six *var*codes circulating in the northwest coast. In this study, as expected comparative analyses to “gold-standard” microsatellite genotyping demonstrated that *var*coding provided higher discriminatory resolution to detect fine-scale genetic variation and describe parasite microevolution related to disease transmission in epidemiological time. These findings were in accordance with the rapidly evolving *var* multigene family due to immune selection^30, 60, 64^, the overall number of microsatellite loci examined and the fact that two loci were fixed in the population^8, 50^ decreasing the denominator of polymorphic microsatellite markers from seven to five. Similar observations have been described in an earlier study in Venezuela where sympatric parasites identical at neutral microsatellite loci were shown to have very different *var* repertoires^65^. These South American genetic epidemiology studies can be compared to those of a peri-urban area in Senegal where reduced transmission has resulted from intense vector and antimalarial control. In Senegal, highly related genotypic clusters of *P. falciparum* isolates were resolved by both *var* and use of a 24-SNP barcode as neutral loci rather than microsatellite loci^66^. Genotyping *var* loci under immune selection in conjunction with neutral loci proves to be informative to validate clonality and avoids the need for whole genome sequencing as a potential rapid response to an outbreak investigation. Even in areas where WGS is routinely conducted, *var*coding has the potential to identify relevant samples for downstream WGS as part of a larger and complementary genomic surveillance framework.

In contrast to the microevolution of parasite *var*codes, we documented conservation of individual *var* DBLα types over space and time. This is similar to Africa^27, 29^ where we see highly variable but many conserved DBLα types yet rapidly evolving *var* repertoires or *var*codes. Here we observed conservation of the outbreak *var*code1 clone DBLα types after transmission within and between multiple human hosts on the scale of months to two years in Ecuador (1,283 independent observations of the same types when examining 36 cases with *var*code1). Moreover, 110 of the 195 (56.4%) Ecuadorian DBLα types were identified across South America over 5 to 13 years (2,130 independent observations of the Ecuadorian DBLα types in 128 different infections from ^28^). These observations are consistent with balancing selection^67^ and in line with other studies demonstrating high levels of *var* DBLα sequence conservation on the time scale of up to 23 years in Brazil^34, 68^. Another independent example of conservation of DBLα sequences over time is found in a peri-urban area of Senegal over 7 years^66^. This pattern of conservation of *var* DBLα types observed in diverse sites confirms the potential of this genetic marker for malaria surveillance.

Of note, the observed conservation of DBLα types in natural infections in humans in Africa^27, 29, 69^ and South America would not be predicted by data from *in vitro* passage of long-term cloned lines of *P. falciparum* showing high rates of mitotic recombination within *var* genes^70–72^. Indeed, Claessens et al ^71^ reported a very high rate of 2-8 x10^-3^ mitotic recombinants created per erythrocytic cycle for three of four long-term *in vitro* cultured lines, “with DBLα domains showing the most recombinations”. The clone tree of HB3, selected for passage through mosquitos, did not show any recombinants suggesting that the high rates observed may relate to passage *in vitro*. While we identified conservation rather than diversification of *var* DBLα types in space and time, it is possible that mitotic recombinants are generated on the observed time scale but not transmitted. Such is the case in HIV (e.g. reviewed in ^73^). Further exploration of within and between host diversification of *var* genes in natural infections requires a more detailed genomic approach analyzing recently adapted parasite lines.

This is the first case study demonstrating the translational application of the *var*code for outbreak surveillance in epidemic/unstable malaria transmission such as E-2020 settings. Specifically, data presented here have shown that *var*coding was able to resolve the population genetics of sustained disease transmission after an outbreak in Ecuador. We conclude that *var*coding requiring a single PCR with degenerate primers has great potential as a simple, cost effective and high-resolution approach to examine *P. falciparum* antigenic diversity in relation to population immunity as well as genome diversification by recombination. This will prove particularly useful in the context of changing patterns of human mobility and gene flow in the Americas where there is high demand for such a diagnostic surveillance method. The tool will also be useful in other epidemic or low transmission settings targeting elimination across the globe.

Here we show that *var* repertoire evolution via sexual recombination was associated with sustained transmission resulting in clinical infections. This is in line with our previous predictions of epidemic disease transmission in South America with parasites with novel *var* repertoires, or *var*codes^28^, and serves as a warning of what may come in terms of monitoring the complexity of malaria resurgence in Latin America with both importation and microevolution of *P. falciparum*. Surveillance with neutral markers alone cannot investigate this potential risk related to recombination of variant antigen gene repertoires. The widespread migration of Venezuelans has already led to massive increases in *P. falciparum* cases in Brazil and Colombia^74^, as well as in the Peru-Ecuador border^3^. Such migration points to the possibility of importation of diverse parasites with antigenically novel *var* genes as we predicted ^28^, and is now also worrisome in the context of increased transmission of malaria in tropical regions due to the diversion of healthcare resources to the COVID-19 pandemic, as recently warned by the WHO^75^. Going the distance to elimination must be supported by appropriate molecular surveillance.

## Methods

### Ethics statement

The *P. falciparum* isolates collected from individuals of all ages presenting with uncomplicated malaria cases were obtained from the malaria surveillance protocol approved by the Ethical Review Committee of Pontificia Universidad Católica del Ecuador. Ethical approval for this study was obtained from Pontificia Universidad Católica del Ecuador (Quito, Ecuador) and University of Melbourne (Melbourne, Australia). Written informed consent was provided by study participants and/or their legal guardians.

### Study design, study area and sample collection

In this molecular epidemiological study, we examined samples that were collected passively from 2013 to 2015 in endemic areas of Ecuador, namely the northwest coast and the Amazon region, as part of ongoing malaria surveillance by the Ecuadorian Ministry of Health and patients showing at the local clinics. During 2013, the majority of samples were collected from an ongoing outbreak that occurred in Esmeraldas city, Esmeraldas province. Details of the sample collection are described in Sáenz et al^8^. In 2014 and 2015, the “post-outbreak” samples were mostly collected from across the northwest coast with few from the Amazon region, reflective of the epidemiology of clinical *P. falciparum* cases in Ecuador (Northwest coast: 20 and 173 cases; Amazon: 11 and 16 cases in 2014 and 2015, respectively). Details of the sample collection are described in Vera-Arias et al ^50^. It is worth noting that although cases were reported throughout 2015, samples were not available for many of them due to logistical challenges. Briefly, subjects who were diagnosed with falciparum malaria either by light microscopy or a rapid diagnostic test were asked if they wanted to participate in the study and signed an informed consent form. The participants had to: 1) live in the areas where the samples were taken, 2) be over 2 years old, 3) agree to participate, 4) sign an informed consent form, 5) give a blood sample, 6) be able to understand the informed consent. From each participant a blood sample (either venous blood or dried blood spot) was collected as well as their responses to a basic demographic questionnaire (including places recently travelled and their address). The positivity of all samples was confirmed in the laboratory by microscopy and PET-PCR ^76^.

### DNA extraction

Genomic DNA was extracted from venous blood or dried blood spot samples using a QIAamp DNA MINI KIT (QIAGEN, Germantown, MD, USA) using the protocol recommended by the manufacturer.

### Microsatellite genotyping

The same *P. falciparum* isolates in this study were previously genotyped for seven putatively neutral microsatellite markers (TA1, POLYα, PfPK2, TA109, 2490, C2M34, C3M39) as described in^8, 50^. These previously published microsatellite genotyping data^8, 48, 75^ were used in the present analyses.

### *Var* genotyping and related data processing

For *var* genotyping, the DBLα domains of antigen-encoding *var* genes were amplified and sequenced on a MiSeq 2×300bp paired-end Illumina platform as described in ^29, 60^. We obtained 6,929,470 raw sequence reads, which we cleaned and processed using the DBLaCleaner pipeline (^60^, http://github.com/Unimelb-Day-Lab/DBLaCleaner). Our customized bioinformatic pipeline has been described in detail in^60^. Briefly, we de-multiplexed and merged the reads as well as removed low-quality reads and chimeras using several filtering parameters (see Supplementary Fig. S1 for details). This resulted in 2,141 cleaned DBLα sequences (Supplementary Fig. S1). To identify distinct or unique DBLα types (i.e., unique genetic variants), we clustered the DBLα sequences from Ecuadorian *P. falciparum* isolates with 5,699 previously published^28^ DBLα sequences from other South American countries (Colombia (N=21 isolates), French Guiana (N=76 isolates), Peru (N=21 isolates), and Venezuela (N=10 isolates) at the standard 96% sequence identity^27^ using the clusterDBLalpha pipeline (http://github.com/Unimelb-Day-Lab/clusterDBLalpha). All the cleaned DBLα sequences in this study have been submitted to the DDBJ/ENA/GenBank (BioProject Number: PRJNA642683).

We further curated our dataset by translating the DBLα types into amino acid sequences using the classifyDBLalpha pipeline (http://github.com/Unimelb-Day-Lab/classifyDBLalpha) and removing any DBLα types that were non-translatable (N=4). In addition, any *P. falciparum* isolate with low sequencing quality (< 10 DBLα types) was removed from the analysis. *Var*coding was completed for a total of 70 *P. falciparum* Ecuadorian isolates, however, 12 *P. falciparum* isolates with low DNA quality and/or poor sequencing quality were removed and we obtained *var* DBLα data for 58 isolates (82.9%). A total of 543 unique DBLα types identified in the 186 South American *P. falciparum* isolates (N = 58 Ecuadorian *P. falciparum* isolates from this study, N = 128 South American *P. falciparum* isolates previously published in ^28^) were used for subsequent *var* analyses at the regional-level, and only the 195 unique DBLα types identified in Ecuador were used for Ecuador-specific analyses.

A further validation of our sequencing and genotyping methodology was undertaken by analyzing the isolate genomic proportions of DBLα types classified as upsA and non-upsA using our classifyDBLalpha pipeline^29^ and are described in Supplementary Text 1.

### Genetic relatedness analyses

#### Measuring pairwise type or allele sharing

To estimate genetic relatedness among isolates, we calculated the similarity index Pairwise Type Sharing (P_TS_) ^26^, as adapted by He et al^60^ to account for differences in *var* DBLα sampling across isolates in the case of *var,* and Pairwise Allele Sharing (P_AS_)^77^ in the case of microsatellites and constructed similarity matrices. Because *P. falciparum* is haploid we refer to *var* repertoires and microsatellite haplotypes in the same manner, i.e. a *var* repertoire of DBLα types or a multilocus haplotype of microsatellite alleles transmitted together. Every parasite isolate was compared to every other parasite isolate in the population to determine the proportion of shared loci (i.e. *var* types or microsatellites), but only “retrospective” pairwise comparisons were analyzed as a conservative approach to account for time order for the Ecuador-specific analyses. In other words, since we were interested in using the outbreak as a baseline this approach allowed us to look at genetic relatedness of an isolate with everything else circulating before its identification. For the comparative analyses with South American data we included all pairwise comparisons and did not correct for time order due to the differences in sampling time points (five to thirteen years before the Ecuadorian isolates were collected).

#### Measuring Bayesian pairwise type sharing

Genetic relatedness (or repertoire overlap) can also be explored more rigorously with unbiased Bayesian pairwise type sharing (BP_TS_, Johnson et al *in preparation*), which accounts for uncertainty in P_TS_ estimates due to differences in isolate repertoire size. This approach uses Bayesian inference methods, which estimate repertoire overlap and uncertainty^78^, and uses them in a subsequent P_TS_ calculation, carrying that uncertainty forward. The prior distribution for repertoire size, used in inference, was informed by observations as follows. First, the median observed repertoire size in Ecuadorian isolates was 37 types, ranging from 11 to 43 (Table S1). Second, the number of expected *var* genes with DBL*α* domains from whole genome sequencing data of the Honduran laboratory reference strain HB3 was 42 ^33^ and 50 *var* genes based on long-read PacBio sequencing of HB3^79^. And third, based on our sequencing data of 37 technical replicates of HB3, the median repertoire size or number of DBL*α* types per isolate was 39 (range 36-41 types), with 40 types consistently identified in the majority of replicates (range 21-37 replicates) (Supplementary Text 1). We therefore used a uniform prior on repertoire sizes between 40 and 50 types, combined with the general Bayesian repertoire overlap framework^78^ to produce unbiased estimates (posterior means). These were used to confirm our P_TS_ estimates. As expected, the P_TS_ and BP_TS_ estimates were positively correlated (Pearson’s correlation coefficient = 0.919, *p* < 0.001, Supplementary Fig. S3). To measure uncertainty in central estimates, we computed a 95% highest density posterior interval (HDPI), a Bayesian version of confidence intervals, for each pairwise estimate. Like a frequentist confidence interval, the width of the HDPI provides a measure of uncertainty of each pairwise comparison. All posteriors were sampled using MCMC.

#### Interpretation of genetic relatedness measures

Since *P. falciparum* undergoes sexual recombination (i.e. meiosis) in the mosquito, conventional genetic interpretations can be applied to P_TS_ and P_AS._ Therefore, pairwise isolate comparisons resulting in a P_TS_ or P_AS_ of 0 (0% shared loci), 0.5 (50% shared loci), and 1 (100% shared loci) would indicate genetically distinct isolates, recombinant isolates, and clones or genetically-identical isolates, respectively. We applied this approach to define “*var*codes” as groups of isolates sharing ≥90% of their DBLα types (P_TS_ ≥ 0.90) to identify putatively identical genomes within the margin of error of detection of 1-5 DBLα types in an isolate (i.e., within the margin of error of detection of all DBLα sequences in an isolate using degenerate primers for *var* DBLα PCR). We confirmed our interpretations of *var*codes, recombinants and genetically distinct isolates by comparing to unbiased BP_TS_ estimates (posterior means) and examining the lower and upper bounds of the 95% HDPIs as a measure of the statistical uncertainty of each pairwise comparison.

In the case of microsatellite genotyping, we applied a threshold of P_AS_ ≥ 0.90 (sharing ≥90% of their microsatellite alleles) to define clones and also applied a threshold of P_AS_ ≥ 0.80 to identify putatively identical genomes within the margin of error of detection of 1 microsatellite allele in an isolate (for comparative purposes to the threshold for *var,* Supplementary Fig. S2). It is important to note that 2 out of the 7 microsatellite markers genotyped were fixed in the population^8^.

#### Visualization of genetic relatedness networks

To visualize the genetic relatedness among isolates as determined by P_TS_ or P_AS_, we constructed networks using the R packages *ggraph*^80^ and *tidygraph*^81^ where isolates are depicted as nodes and edges as the P_TS_ or P_AS_ values at a given threshold. The R package *ggspatial*^82^ was used to plot spatial networks using latitude/longitude coordinates for sampling location. To visualize genetic relatedness of parasites over time we used the R package *gganimate*^83^ to construct spatiotemporal relatedness networks. We generated a clustered heatmap based on the presence/absence matrix of DBLα types to visualize the genetic profiles of each isolate in Ecuador and each country in South America using the R package *pheatmap*^84^ and the “complete” clustering method. Unrooted neighbor-joining phylogenetic trees based on pairwise genetic distance (calculated as 1-P_TS_) were constructed using the R packages *ape*^85^ and *ggtree*^86, 87^.

### Statistical analysis

We used R version 3.5.2 for all analyses^88^. The package *dplyr*^89^ was used for further data curating. We used base R and the R package *epiR*^90^ to perform chi-squared tests for univariate analyses of categorical variables to compare proportions and for non-parametric tests to compare distributions of continuous variables between two groups (Mann-Whitney U test) or among *k* groups (Kruskal-Wallis test) with a Bonferroni correction for multiple comparisons. To evaluate how well we sampled the true pool of *var* DBLα diversity (i.e. the true number of genetic variants circulating) in Ecuador and in South America, we used the R package *vegan*^91^ to generate species accumulation curves.

### Data availability

The cleaned DBLα sequences generated in this study from Ecuadorian *P. falciparum* isolates have been submitted to the DDBJ/ENA/GenBank (BioProject Number: PRJNA642683). The python code for the DBLαCleaner pipeline is available on GitHub at https://github.com/Unimelb-Day-Lab/DBLaCleaner. The python code for the clusterDBLalpha pipeline is available on GitHub at https://github.com/Unimelb-Day-Lab/DBLalpha. The python code for the classifyDBLalpha pipeline is available on GitHub at https://github.com/Unimelb-Day-Lab/classifyDBLalpha. All other deidentified data and analysis code are available on the open-source GitHub repository at https://github.com/shaziaruybal/varcode-ecuador.

## Data Availability

The cleaned DBLα sequences generated in this study from Ecuadorian P. falciparum isolates have been submitted to the DDBJ/ENA/GenBank (BioProject Number: PRJNA642683). The python code for the DBLαCleaner pipeline is available on GitHub at https://github.com/Unimelb-Day-Lab/DBLaCleaner. The python code for the clusterDBLalpha pipeline is available on GitHub at https://github.com/Unimelb-Day-Lab/DBLalpha. The python code for the classifyDBLalpha pipeline is available on GitHub at https://github.com/Unimelb-Day-Lab/classifyDBLalpha. All other deidentified data and analysis code are available on the open-source GitHub repository at https://github.com/shaziaruybal/varcode-ecuador.

## Acknowledgements

The authors would like to thank the patients who contributed samples and the field teams who were involved in sample collections. We thank the Ecuadorian Ministry of Health, especially Luis E Castro, Julio Valencia and Javier Gomez Obando. We appreciate the support of the Australian Genome Research Facility for illumina sequencing of the samples. This research was financially supported by the Pontificia Universidad Católica del Ecuador (grant numbers: L13058, L13248, M13416, N13416, O13087[QINV0084] to F.E.S), the Fogarty International Center at the National Institutes of Health [Program on the Ecology and Evolution of Infectious Diseases (EEID), Grant number: R01-TW009670 to K.P.D.], and the National Institutes of Allergy and Infectious Diseases, National Institutes of Health (grant number: R01-AI084156 to K.P.D.). S.R-P. was supported by a Melbourne International Engagement Award from the University of Melbourne and gratefully acknowledges the J.D. Smyth Travel Award from the Australian Society for Parasitology that enabled her to travel to Ecuador to establish this collaborative project.

## Author contributions

S.R-P. and K.P.D. conceived and designed the study; F.E.S. and C.A.V-A. carried out field work to obtain *P. falciparum* isolates and epidemiological metadata; S.R-P. and K.E.T. developed the *var*coding methodology; S.R-P. and S.D. performed the molecular experiments and sequencing; S.R-P., F.E.S. and C.A.V-A. curated the clinical and epidemiological metadata; S.R-P. performed the formal data analysis and visualization; E.K.J. and D.B.L. designed and conducted the Bayesian statistical analysis; S.R-P., F.E.S., K.E.T. and K.P.D. contributed to the interpretation of the data; K.P.D. supervised the work; S.R-P., F.E.S. and K.P.D. acquired funding; S.R-P. and K.P.D. wrote the paper with contributions from all authors.

## Supplementary Information

### Supplementary Figures

**Figure S1.**
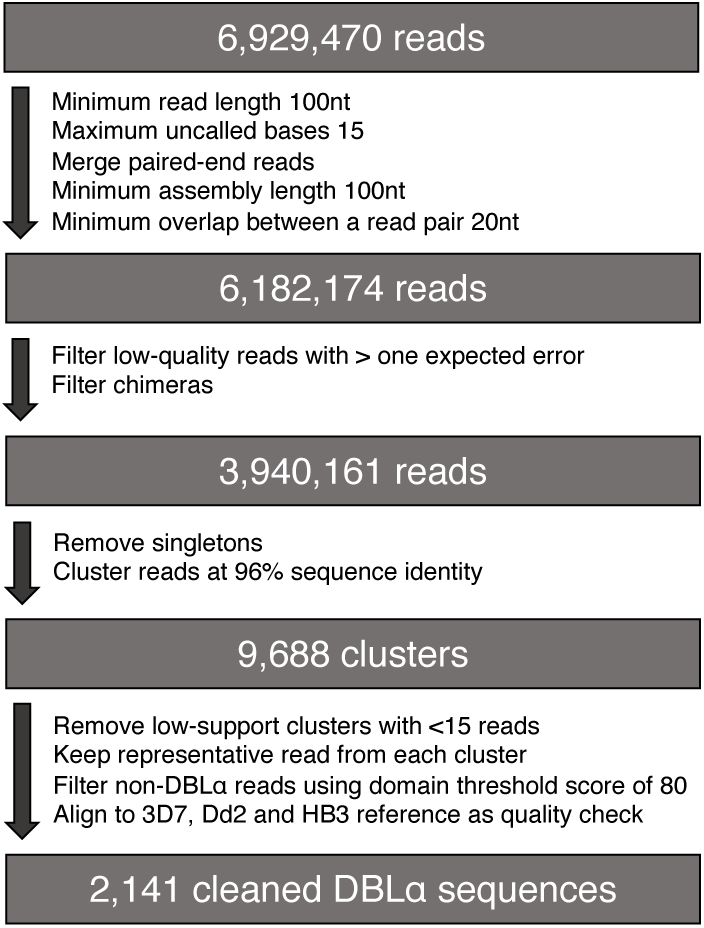
Bioinformatic sequence data processing flowchart. The flowchart shows the bioinformatic process to clean the raw illumina sequence reads with details on filter parameters for each step. This custom bioinformatic pipeline was developed to de-multiplex and remove PCR and sequencing artefacts and is described in detail in He et al ^60^.

**Figure S2.**
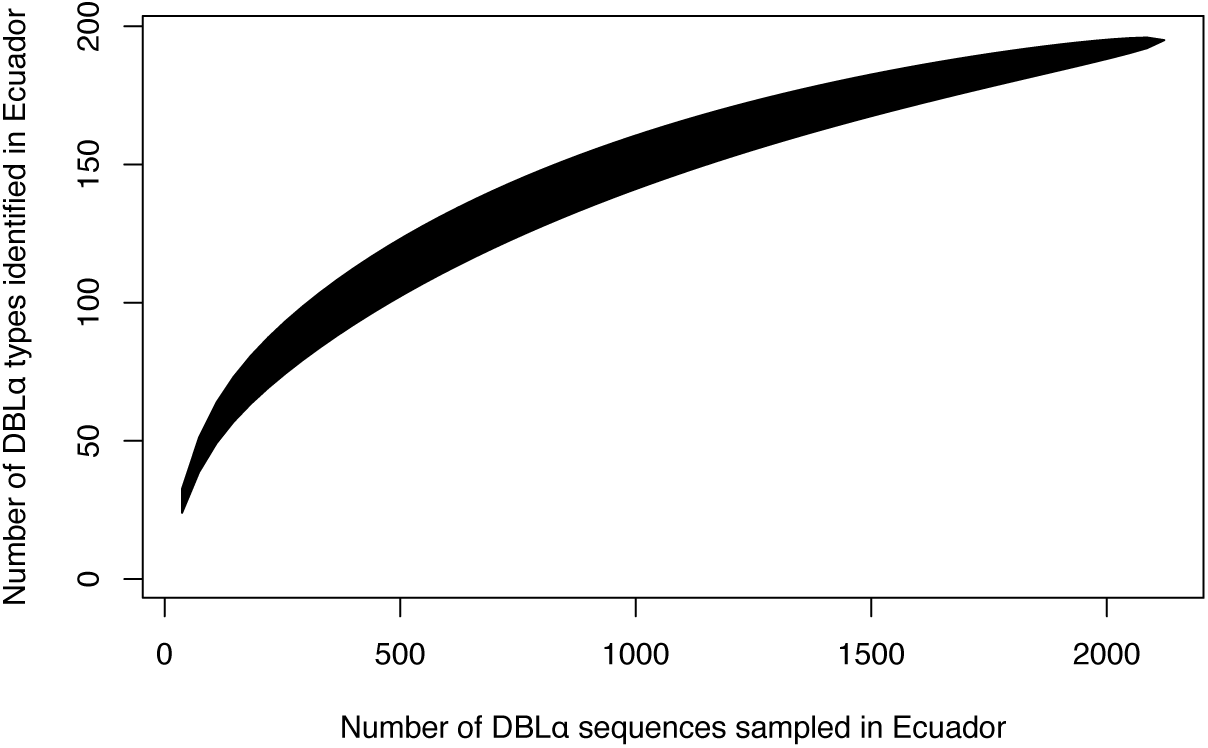
Sampling depth of *var* DBLα types in Ecuador. Accumulation curves depict the number of observed DBLα types plotted as a function of the number of DBLα sequences sampled.

**Figure S3.**
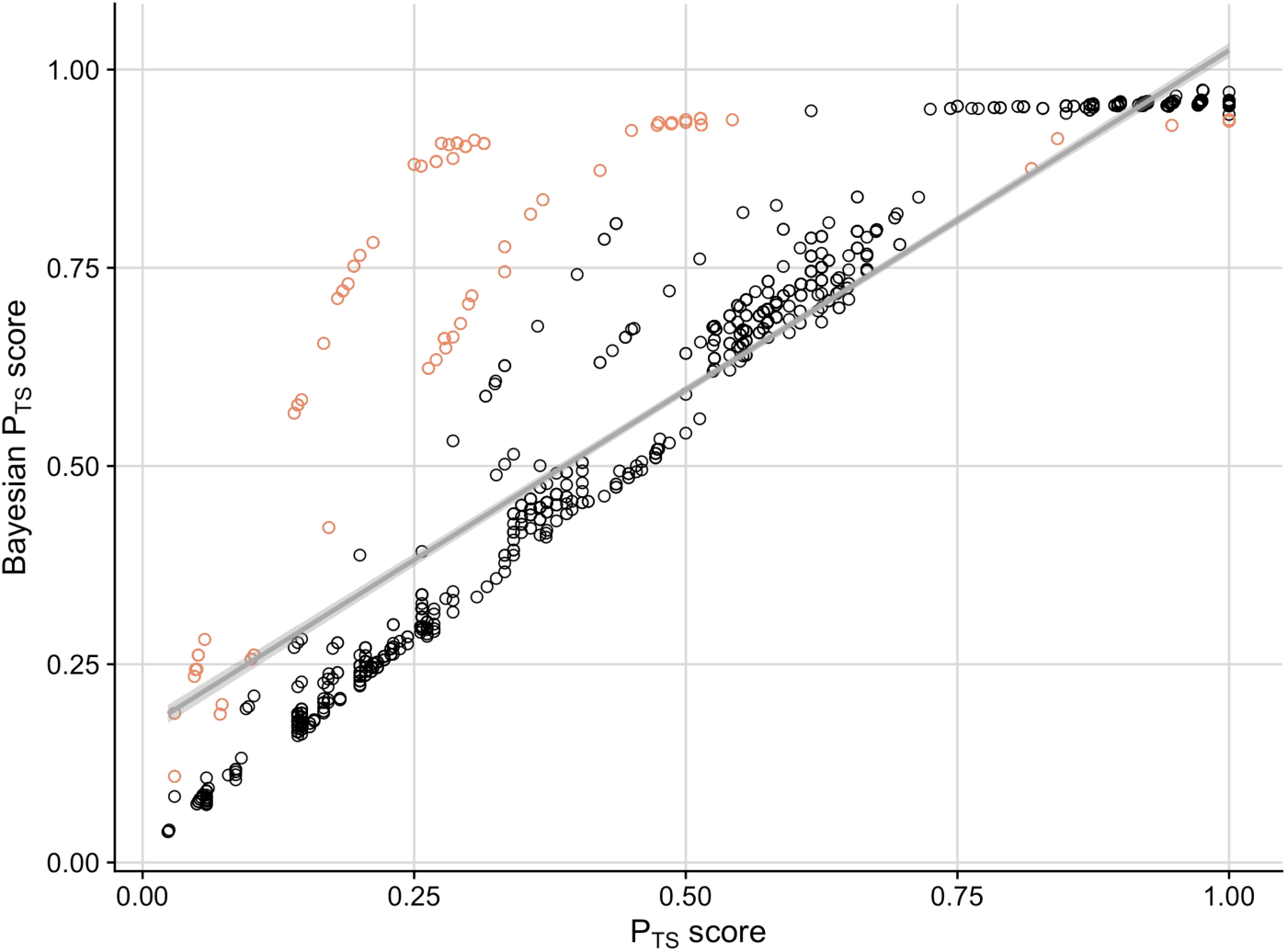
Relatedness of isolates based on observed and unbiased Bayesian pairwise type sharing estimates. There was a positive correlation between the two relatedness measures (correlation coefficient = 0.919, *p* < 0.001). Circles colored in light red correspond to pairwise comparisons involving the two *P. falciparum* isolates with the smallest repertoire sizes (11 and 19 DBLα types), many of which had lower statistical confidence and the largest discrepancies between observed and unbiased estimates.

**Figure S4.**
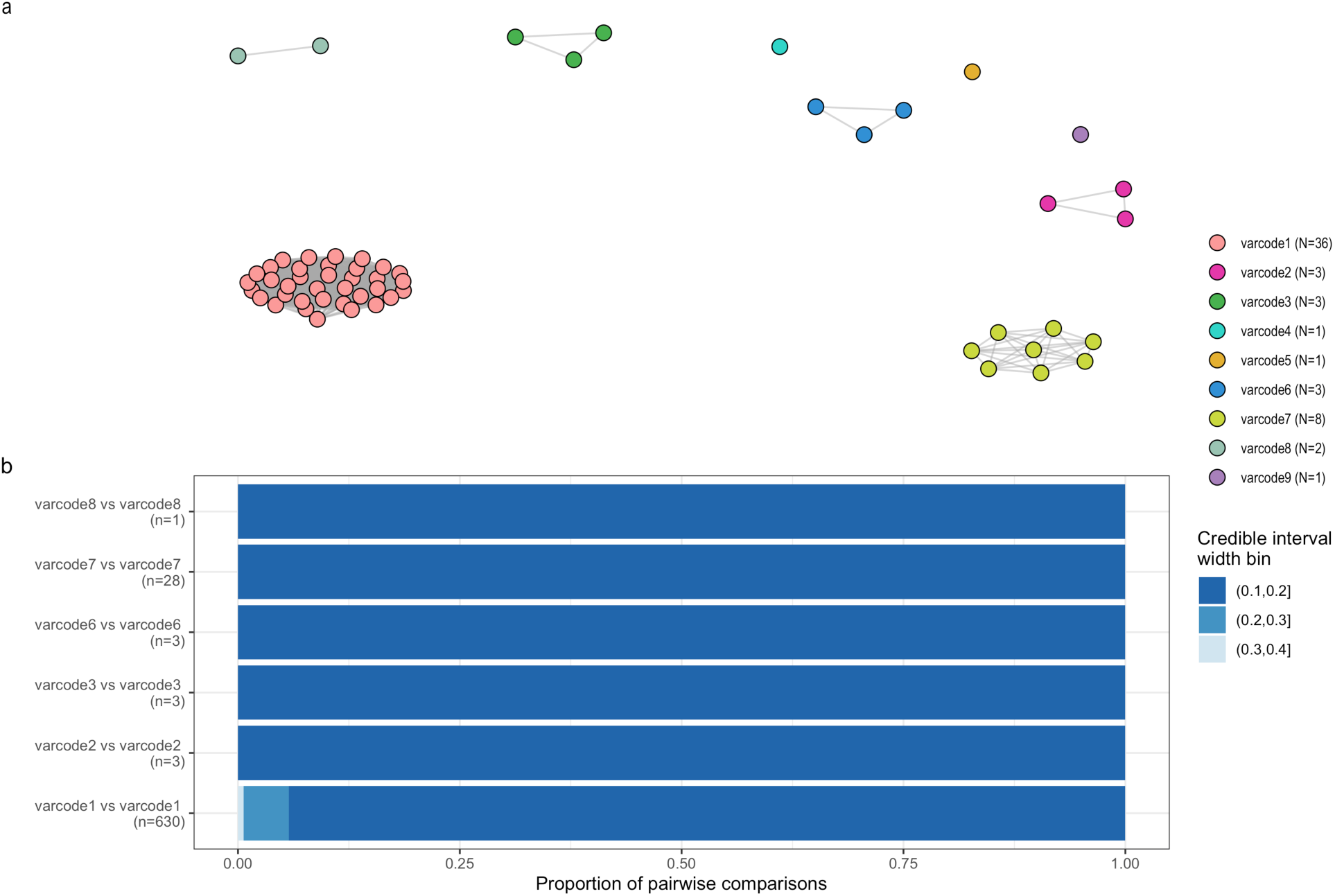
Genetic relatedness networks based on Bayesian pairwise type sharing estimates to define *var*codes. A network visualization of the genetic relatedness of *P. falciparum* isolates at the threshold of (a) BP_TS_ ≥0.90 to define clusters of genetically-related isolates based the mean posterior Bayesian PTS (BP_TS_). Every node represents a *P. falciparum* isolate and an edge represents the BP_TS_ value between two particular nodes/isolates. Isolates that cluster together (i.e., connected by edges) are considered to be genetically identical (i.e., clones). Every color represents a different *var*code. (b) The proportion of pairwise comparisons within a given 95% highest density posterior interval width, which provides a measure of the uncertainty of each pairwise estimate. Only *var*codes with >1 *P. falciparum* isolate are shown. For the full distribution of within-*var*code BP_TS_ estimates see Fig. S4b.

**Figure S5.**
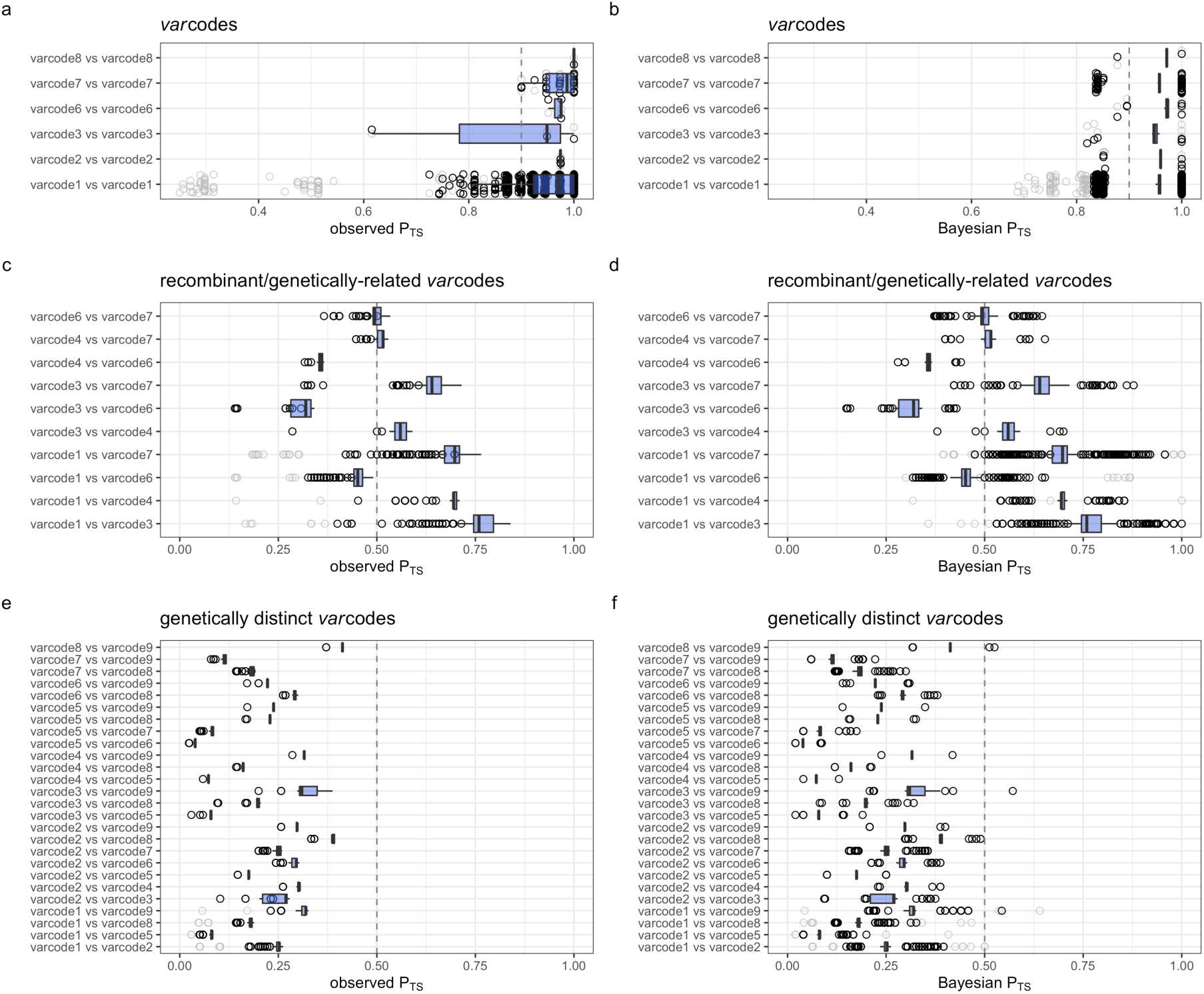
Distribution of observed and Bayesian pairwise type sharing estimates used to define *var*codes, as well as recombinant, genetically-related, and genetically distinct *var*codes. For observed P_TS_ estimates (a, c, e), the boxplots (in blue) show the median and interquartile range of the observed P_TS_ estimates (circles) for all pairwise comparisons among *P. falciparum* isolates for a given within-*var*code or between-*var*code comparison. For Bayesian PTS (BP_TS_) estimates (b, d, f), the boxplots (in blue) show the median and interquartile range of the posterior mean BP_TS_ estimates. The circles show the BP_TS_ estimates for the lower and upper bound of the 95% highest density posterior intervals (HDPIs). Circles showing all of the posterior mean BP_TS_ estimates are omitted for clarity. Dashed lines indicate the threshold of (a-b) P_TS_ or BP_TS_ ≥0.90 and (c-f) P_TS_ or BP_TS_ ≥0.50. For comparison to the genetic relatedness network edges see Figures 2a, 2c, S3a, S7a. In all panels, light grey circles correspond to pairwise comparisons involving the two *P. falciparum* isolates with the smallest repertoire sizes (11 and 19 DBLα types), many of which had lower statistical confidence and the largest discrepancies between observed and unbiased estimates. Black circles denote all other estimates. Only *var*codes with >1 *P. falciparum* isolate are shown.

**Figure S6.**
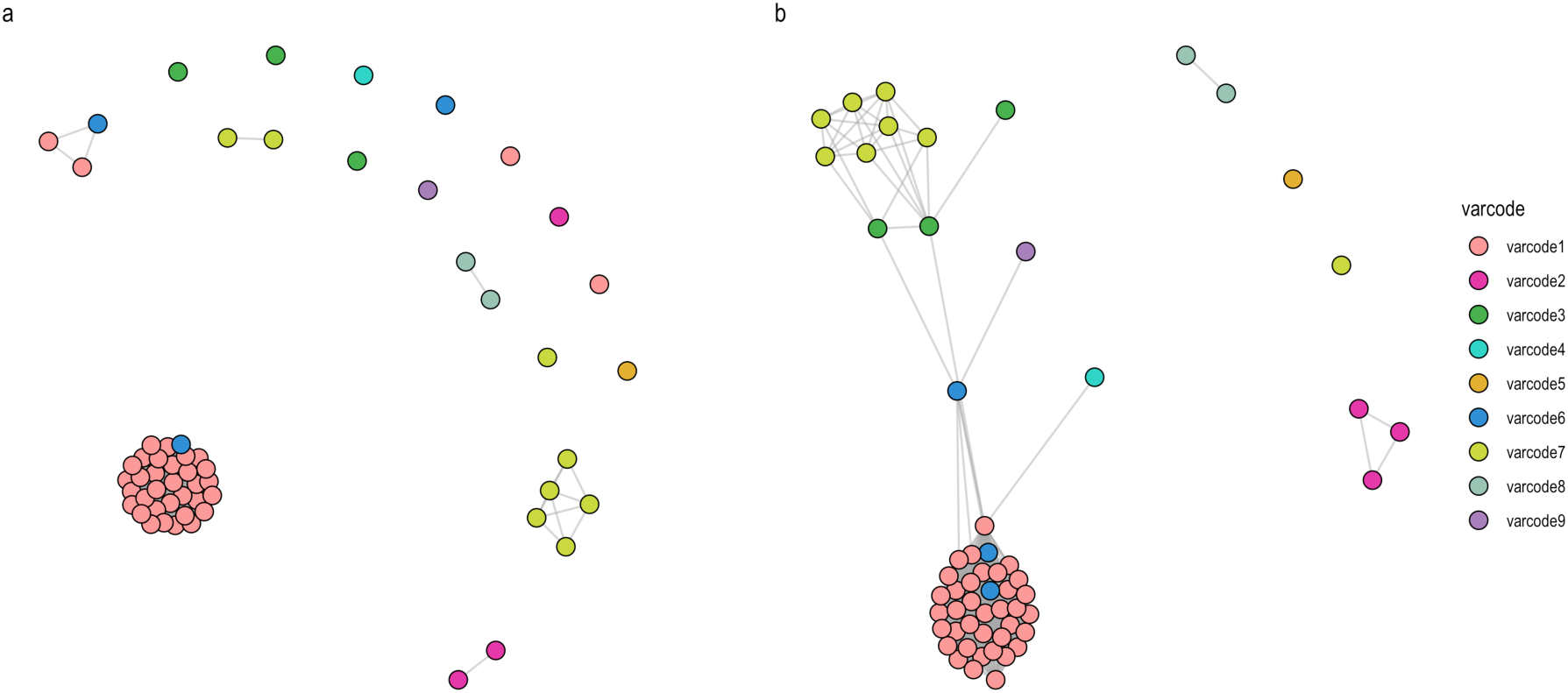
Genetic relatedness networks by microsatellite genotyping. A network visualization of the genetic relatedness of *P. falciparum* isolates at the threshold of (a) P_AS_ ≥0.90 and (b) P_AS_ ≥0.80 to define clusters of genetically-related isolates based on microsatellite genotyping. Every node represents a *P. falciparum* isolate and an edge represents the P_AS_ value between two particular nodes/isolates. Isolates that cluster together (i.e., connected by edges) are considered to be genetically identical (i.e., clones) at the threshold of P_AS_ ≥0.90 in (a) but can differ by 1 allele at the threshold of P_AS_ ≥0.80 in (b). Every color represents a different *var*code and are colored based on the definition of *var*codes by *var* for comparative purposes. It is important to note that 2 out of the 7 microsatellite markers genotyped were fixed in the population^8^.

**Figure S7.**
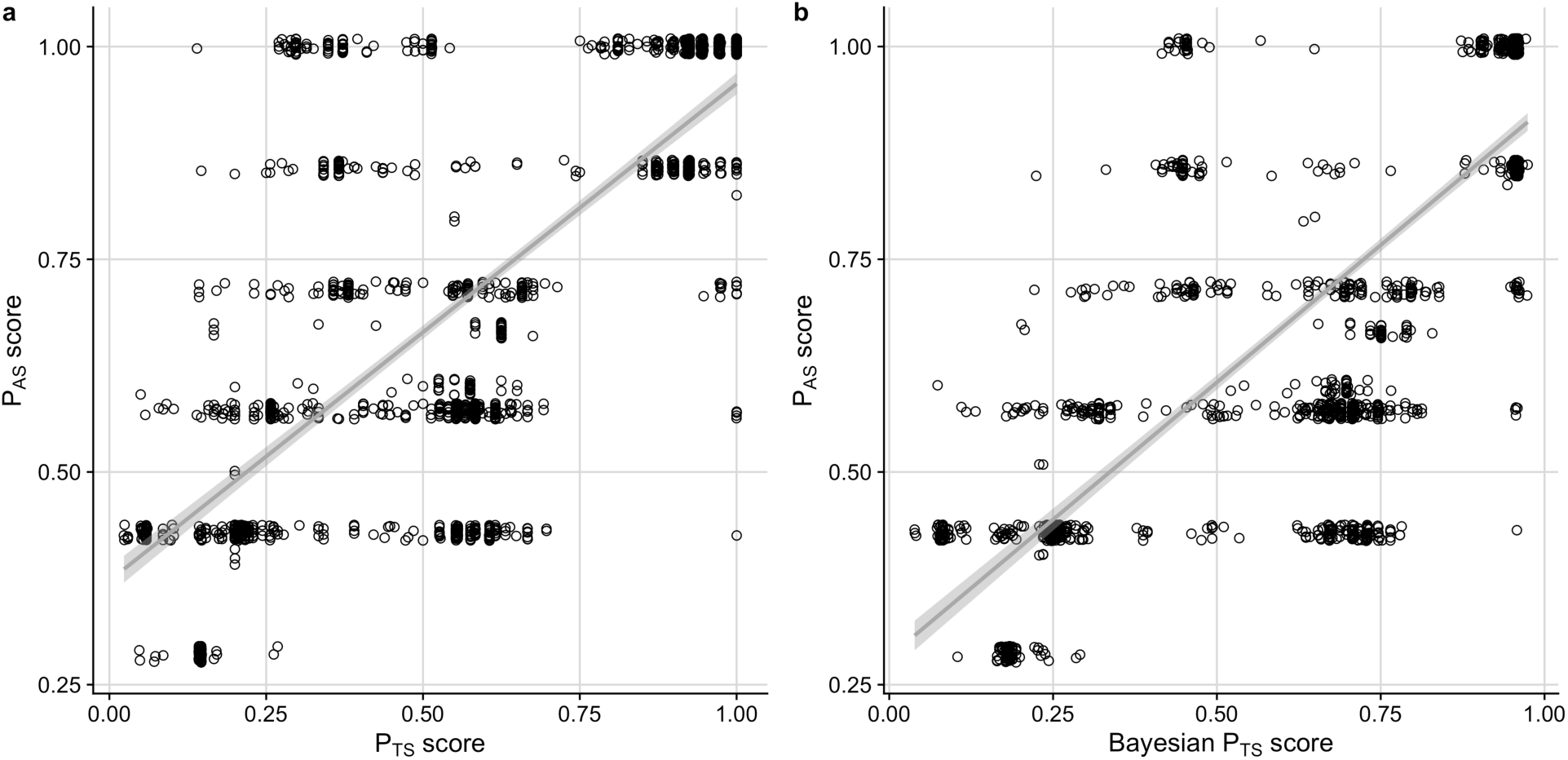
Relatedness of isolates based on *var* and microsatellite genotyping. There was a positive correlation between (a) P_TS_ and P_AS_ relatedness measures (correlation coefficient = 0.757, *p* < 0.001) as well as (b) unbiased Bayesian P_TS_ and P_AS_ relatedness measures (correlation coefficient = 0.779, *p* < 0.001).

**Figure S8.**
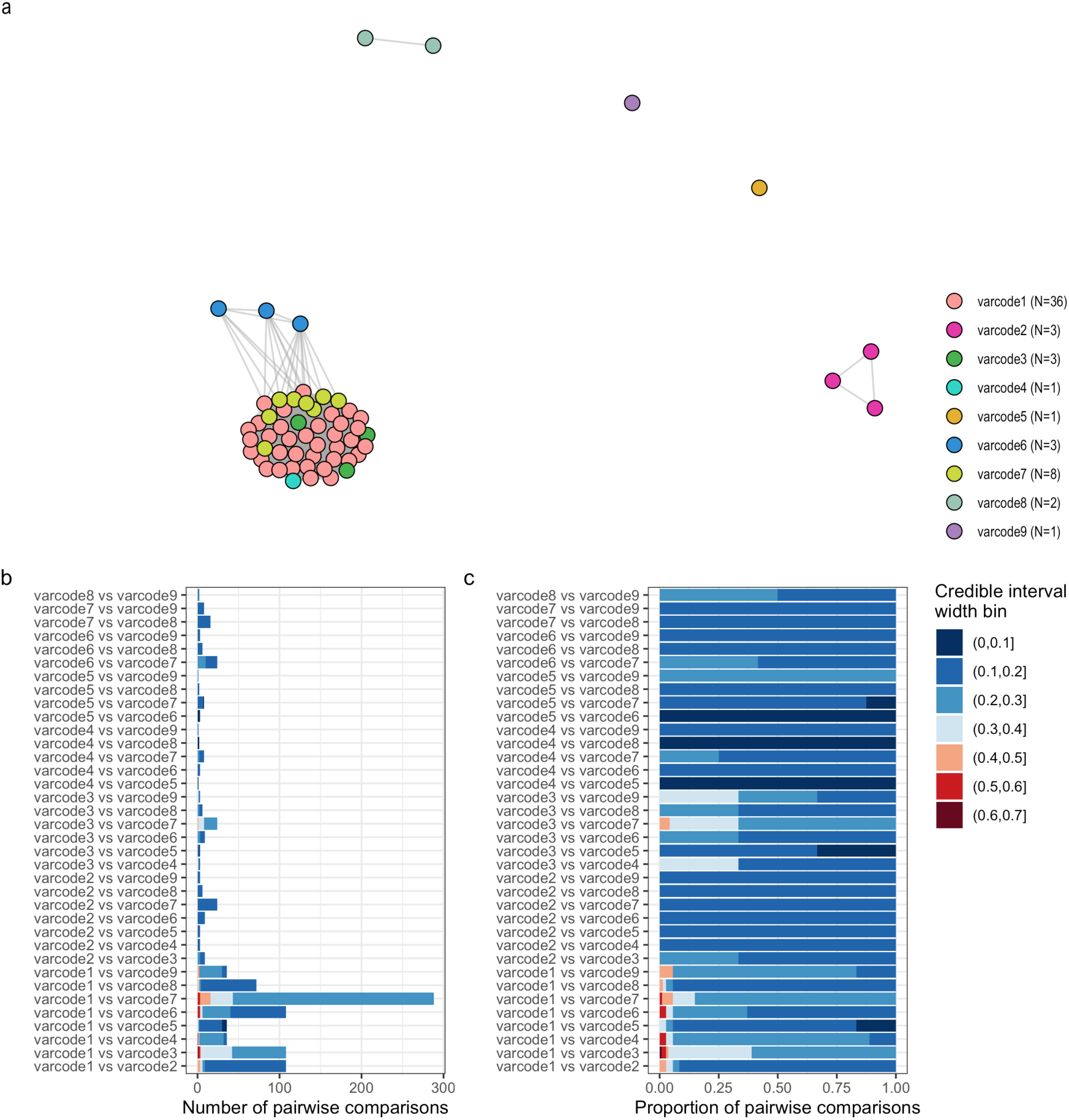
Genetic relatedness networks based on unbiased Bayesian pairwise type sharing estimates to identify recombinants. A network visualization of the genetic relatedness of *P. falciparum* isolates at the threshold of (a) BP_TS_ ≥0.50 to define clusters of genetically-related isolates based on the posterior mean BP_TS_ estimates. Every node represents a *P. falciparum* isolate and an edge represents the BP_TS_ value between two particular nodes/isolates. Isolates that cluster together (i.e., connected by edges) are considered to be genetically identical (i.e., clones). Every color represents a different *var*code. The (b) number and (c) proportion of pairwise comparisons within a given 95% highest density posterior interval width, which provides a measure of the uncertainty of each pairwise estimate. For the full distribution of within-*var*code BP_TS_ estimates see Fig. S4d-f.

**Figure S9.**
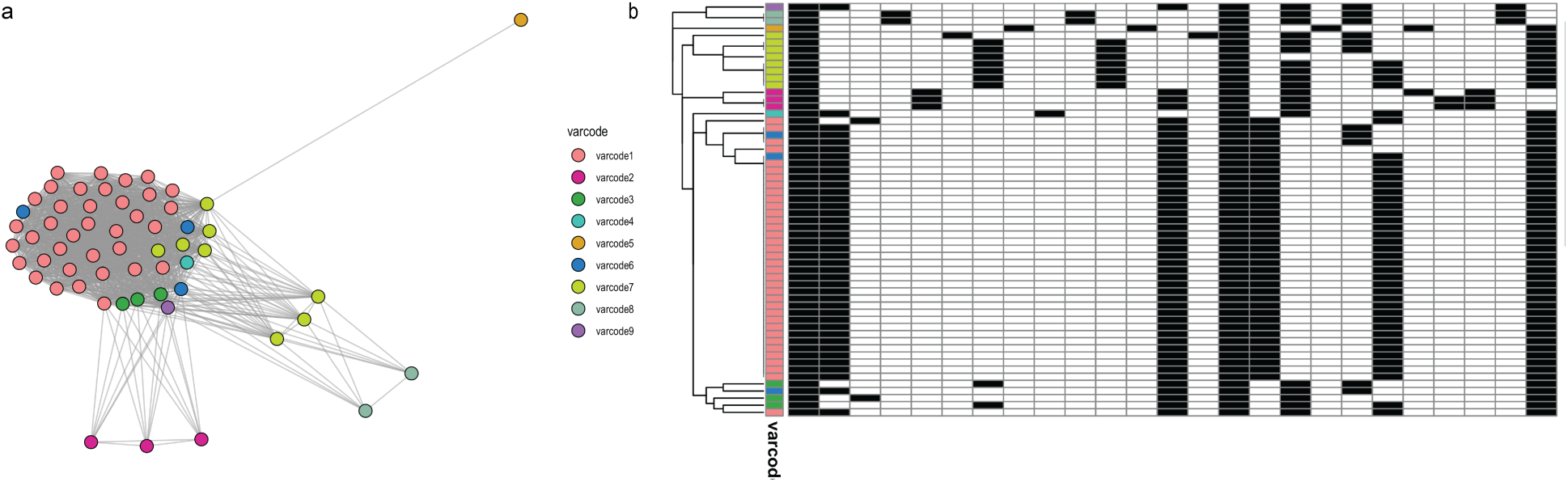
Relatedness networks based on microsatellite genotyping. (a) A network visualization of the genetic relatedness of *P. falciparum* isolates at the threshold of P_AS_ ≥0.50 colored by *var*code for comparative purposes. Every node represents a *P. falciparum* isolate and an edge represents the P_AS_ value between two particular nodes/isolates. (b) A clustered heatmap showing the genetic profiles of each *P. falciparum* isolate with rows representing each isolate and columns representing each microsatellite allele. Black and white denote the presence and absence of each allele, respectively. Isolates that clustered together were more genetically similar (i.e., the same types were present).

**Figure S10.**
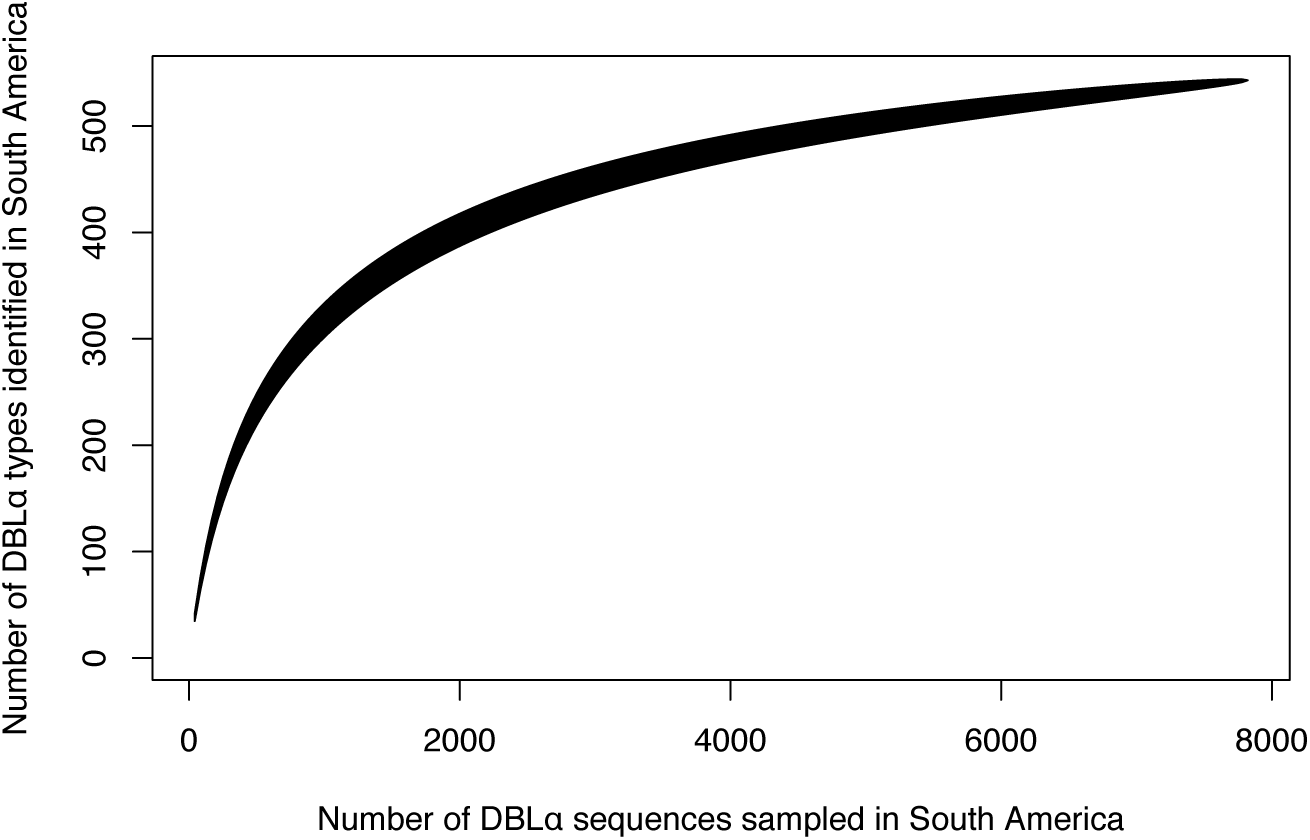
Sampling depth of *var* DBLα types in South America. Accumulation curves depict the number of observed DBLα types plotted as a function of the number of DBLα sequences sampled.

**Figure S11.**
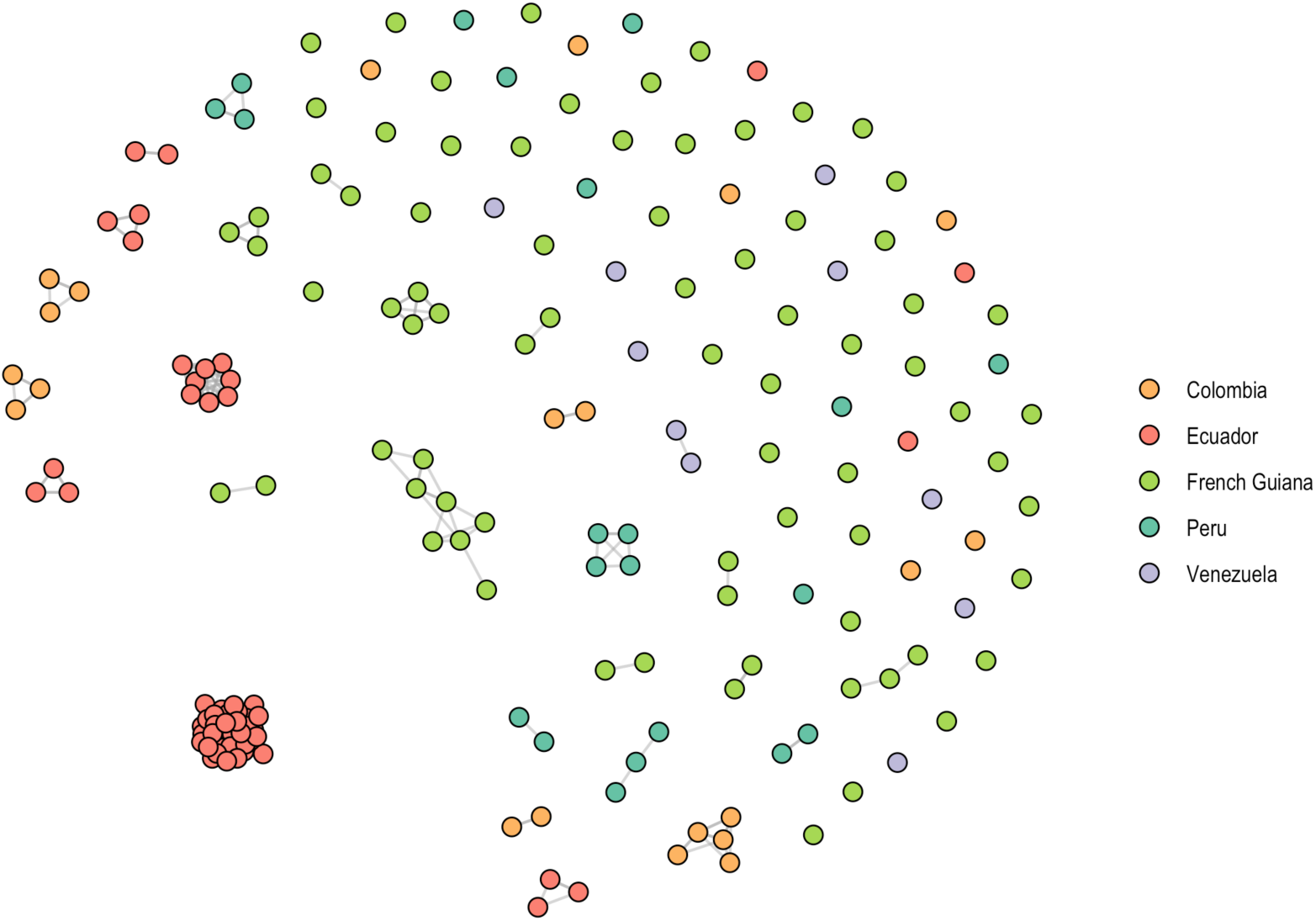
South American *var*codes. A network visualization of the genetic relatedness of *P. falciparum* isolates at the threshold of P_TS_ ≥0.90 to define clusters of genetically-related isolates, or *var*codes in South America. Every node represents a *P. falciparum* isolate and an edge represents the P_TS_ value between two particular nodes/isolates. Isolates that cluster together (i.e., connected by edges) are considered to have the same *var*code. The colors depict the country of origin/sampling location of each *P. falciparum* isolate.

**Figure S12.**
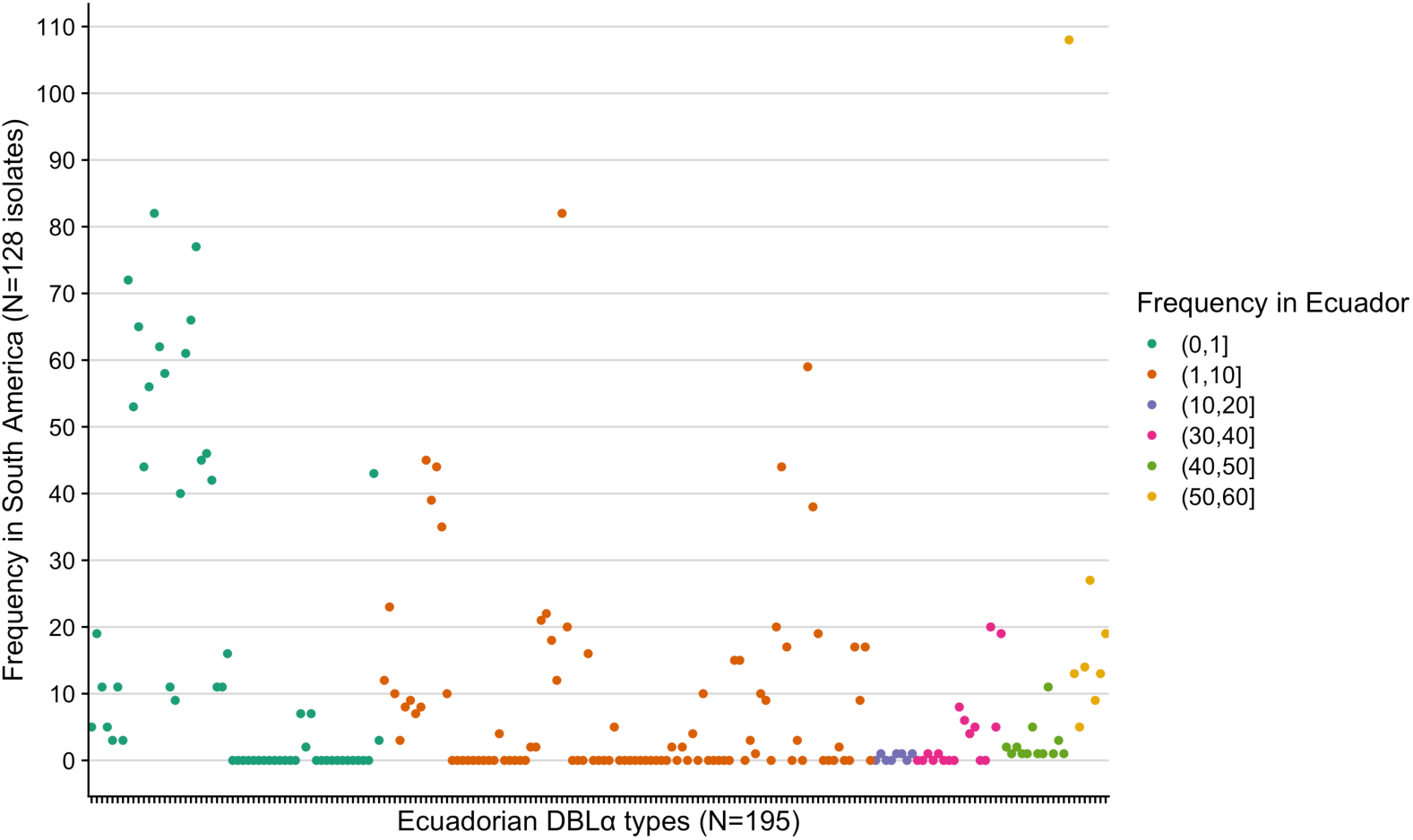
Frequency distribution of the 195 DBLα types identified in Ecuador. Each point corresponds to an individual DBLα type and is colored by its frequency (i.e., the number of Ecuadorian *P. falciparum* isolates it was identified in). The y-axis depicts the frequency at which each Ecuadorian DBLα type was identified in the 128 *P. falciparum* isolates from South America. Of the 56 “singleton” DBLα types (i.e., frequency of 1) in the Ecuadorian dataset, 32 (57.1%) were identified in other South American countries and 24 (42.9%) were not identified anywhere else in South America.

**Figure S13.**
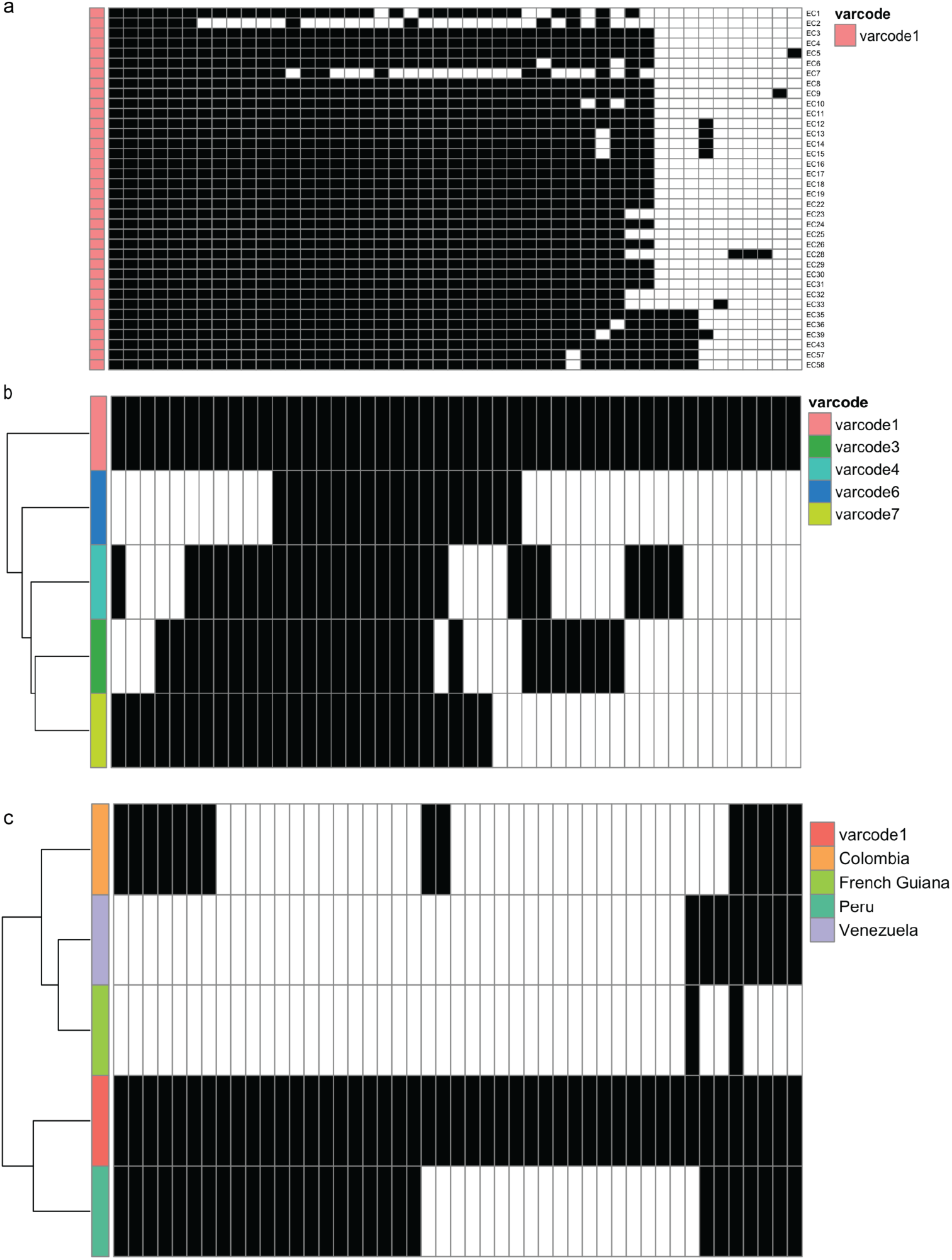
Identification of the outbreak *var*code1 DBLα types. Each column represents one of the 47 DBLα types and black and white denote the presence and absence of each type, respectively, in (a) the 36 infections with *var*code1, (b) in the 15 infections with recombinant *var*codes and (c) in South American *P. falciparum* populations. (a) The rows represent case numbers and are not clustered such that from top to bottom they are representative of sampling date (see Table 1 for details on sampling dates and whether they correspond to during or post-outbreak), and the columns are ordered from left to right in decreasing frequency of each DBLα type. *Note*: EC2 and EC7 were under sampled as compared to the other *P. falciparum* isolates (11 and 19 DBLα types per isolate, respectively. (b-c) Clustered heatmaps show the identification of the 47 DBLα types from the outbreak *var*code1 *P. falciparum* isolates in the other *P. falciparum* isolates from South America. A total of 31 (66%) of the outbreak types were identified in one or more countries. The highest number of types identified in another country was 28 (60%) in isolates collected in 2003-2004 from Peru, followed by 14 (30%) in isolates collected from 2002-2006 from Colombia, followed by 8 (17%) in isolates collected from 2003-2007 from Venezuela and only 2 (4%) in isolates collected from 2006-2008 from French Guiana. The isolate collection dates are reported in^28^.

### Supplementary Tables

**Table S1.** Summary statistics for total number of *var* DBLα types per isolate (i.e., repertoire size) in South American *P. falciparum* populations.

| Location | min* | med | mean | max |
| --- | --- | --- | --- | --- |
| Colombia | 19.0 | 40.0 | 38.4 | 43.0 |
| Ecuador | 11.0 | 37.0 | 36.6 | 43.0 |
| French Guiana | 13.0 | 48.0 | 50.5 | 92.0 |
| Peru | 19.0 | 36.0 | 33.4 | 42.0 |
| Venezuela | 28.0 | 36.5 | 35.2 | 43.0 |

\*Note: all isolates with <10 types were removed due to low sequencing quality (see Methods)

## Supplementary Text 1

As described in the Methods, the degenerate primers we use amplify the DBL*α* domain of *var* genes. We performed an additional validation of our PCR (using these degenerate DBL*α* primers) and sequencing methodology to understand the margin of error of detection of all DBL*α* types in an isolate. DBL*α* types were translated into amino acid sequences and classified as upsA or non-upsA, using the classifyDBLalpha pipeline (Ruybal-Pesántez et al. 2017) to examine whether the expected genomic proportions of upsA/non-upsA were obtained in each isolate.

Given our study was conducted in South America, we used the Honduran laboratory reference strain HB3 as a benchmark for the expected number of total DBL*α* types (i.e. repertoire size), as well as the proportion of upsA/non-upsA. Whole genome sequencing of HB3 has identified 44 *var* genes, with 8 upsA DBL*α* types (defined by DBL*α* domain 1), 34 non-upsA DBL*α* types (DBL*α* domains 0 and 2) and 2 upsE (*var2csa* genes, defined by DBLpam domains and do not have DBL*α* domains) (Rask et al. 2010). We independently verified this using 37 technical replicates of *var* DBL*α* PCR amplification and illumina sequencing of our HB3 laboratory isolate. It is worth noting that since HB3 has two *var2csa* genes that do not have DBL*α* domains, these will not be amplified by our degenerate primers, so we expect that only 42 of the *var* genes of HB3 could be amplified.

### HB3 technical replicates

From the data obtained from 37 HB3 technical replicates, we identified the expected repertoire sizes with a median of 39 DBL*α* types (range: 36-41). A median of 7 upsA DBL*α* types (range: 6-8) and a median of 33 non-upsA DBL*α* types (range: 30-34) were identified (Figure 1). The median genomic proportion of upsA DBL*α* types was 17.5% (range: 15.4-19.5%) and 82.5% (range: 80.5-84.6%) for non-upsA DBL*α* types (Figure 2). We found that of the 46 identified in the 37 technical replicates, 40 of them were consistently identified in the majority of replicate isolates (range 21 to 37 replicates). Of these 40 types, 7 were ups-A and 33 were non-upsA. All of these findings are in line with what is expected from whole genome sequencing data (Rask et al. 2010).

**Figure 1:**
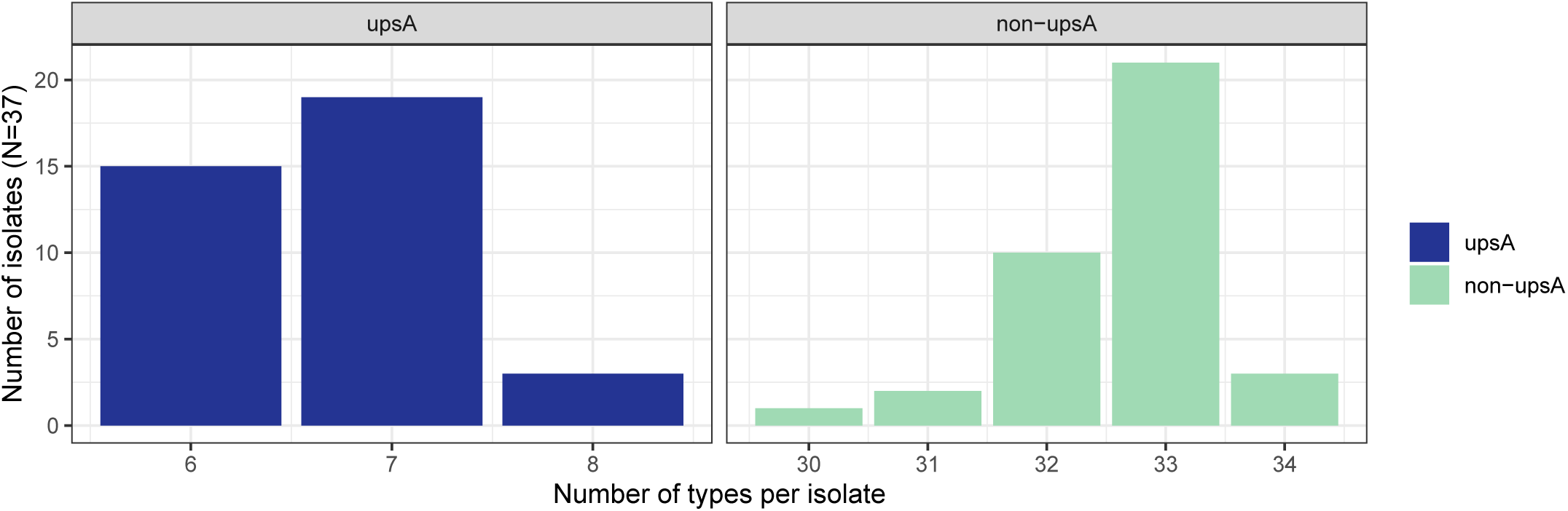
The distribution of the number of upsA and non-upsA types identified in each HB3 isolate repertoire.

**Figure 2:**
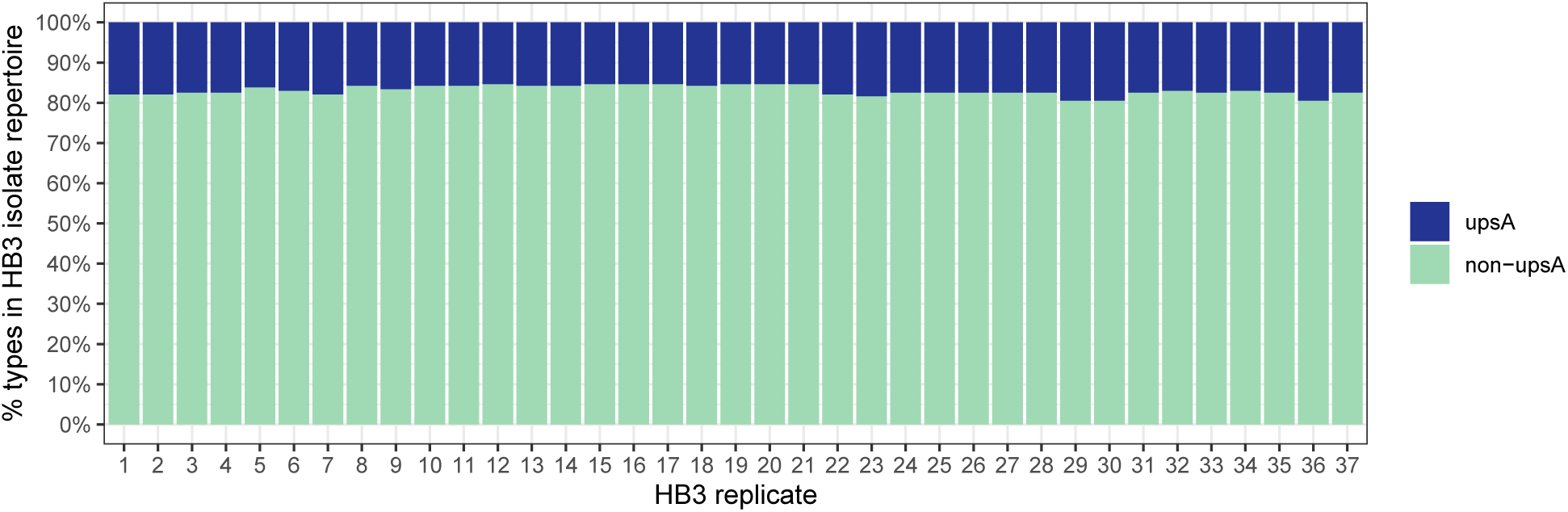
The proportion of upsA and non-upsA types identified in each HB3 isolate repertoire.

### Field isolate genomic proportions of upsA and non-upsA DBL*α* types

The 543 unique DBL*α* types identified in the 186 South American *P. falciparum* isolates (N = 58 Ecuadorian P. falciparum isolates from this study, N = 128 previously published *P. falciparum* isolates from Colombia, French Guiana, Peru and Venezuela) were translated into amino acid sequences and classified as upsA or non-upsA, using the classifyDBLalpha pipeline (Ruybal-Pesántez et al. 2017). There were 79 upsA and 464 non-upsA types.

Looking first at the 195 types (26 upsA and 169 non-upsA) identified in Ecuadorian isolates, we obtained a median genomic proportion of upsA of 10.8% (range: 5.1-18.2%) and 89.2% (range: 81.8-94.9%) of non-upsA types for all isolate *var* codes (Figure 3).

**Figure 3:**
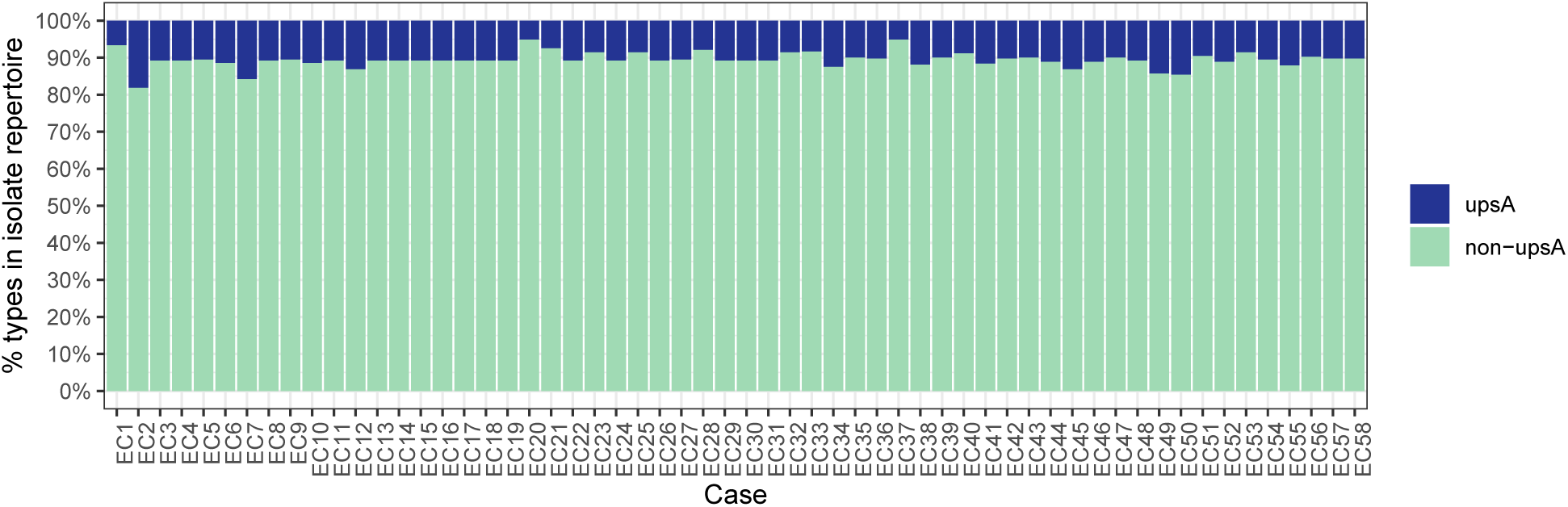
The proportion of upsA and non-upsA types in each isolate repertoire. The case numbers in the x-axis can be used to identify the clinical information of the participant in Table 1.

To confirm the patterns we observed in Ecuadorian isolates, we also compared them to the other South American isolates. In the other South American isolate *var* codes the genomic proportions of upsA/non-upsA were similar, with a median proportion of upsA of 9-14% and 86-91% for non-upsA types (Table 1).

**Table 1:** The proportion of upsA types in South American isolates

| country | min | median | mean | max |
| --- | --- | --- | --- | --- |
| Colombia | 0.00 | 0.09 | 0.09 | 0.14 |
| Ecuador | 0.05 | 0.11 | 0.11 | 0.18 |
| French Guiana | 0.04 | 0.11 | 0.11 | 0.18 |
| Peru | 0.08 | 0.14 | 0.14 | 0.20 |
| Venezuela | 0.05 | 0.11 | 0.12 | 0.21 |

With regards to the number of upsA identified in all the South American isolate *var* codes, we identified a median of 4-5 upsA types and 31-42.5 non-upsA types (Table 2).

**Table 2:** The number of upsA types identified in South American isolates

| country | min | median | mean | max |
| --- | --- | --- | --- | --- |
| Colombia | 0 | 4 | 4 | 6 |
| Ecuador | 2 | 4 | 4 | 6 |
| French Guiana | 2 | 5 | 6 | 14 |
| Peru | 2 | 5 | 5 | 7 |
| Venezuela | 2 | 4 | 4 | 9 |

### Inheritance of DBL*α* types in the recombinant *var*codes

For the outbreak *var* code1, its 47 DBL*α* types were classified as 5 upsA and 42 non-upsA. In Figure 5 we look specifically at the “inheritance” of upsA (blues) vs non-upsA (greens) types from the parental outbreak *varc*ode1 in the case of the parasites with recombinant *var* codes (*var* codes3,4,6,7). This provides a proxy to examine inheritance of types with regards to their chromosomal location. The proportion of the outbreak types that were inherited in the recombinant parasite *var* codes is indicated in the darker shades of blue or green, showing that the proportion of inherited types varied both by *var* code and upsA/non-upsA. Overall, the DBL*α* type sharing patterns in parasites with recombinant varcodes are consistent with inheritance of 50% types, with a higher proportion of non-upsA inherited types (∼40-70%, i.e. 17 to 30 of the 42 types) vs upsA (∼20-50%, i.e. 1 to 3 of the 5 upsA types). The exception was *var* code7 where 60-80% of upsA were inherited, i.e. 3-4 of the 5 upsA types. The lighter shades correspond to those types that were not inherited from the outbreak clone but from the other parent.

**Figure 4:**
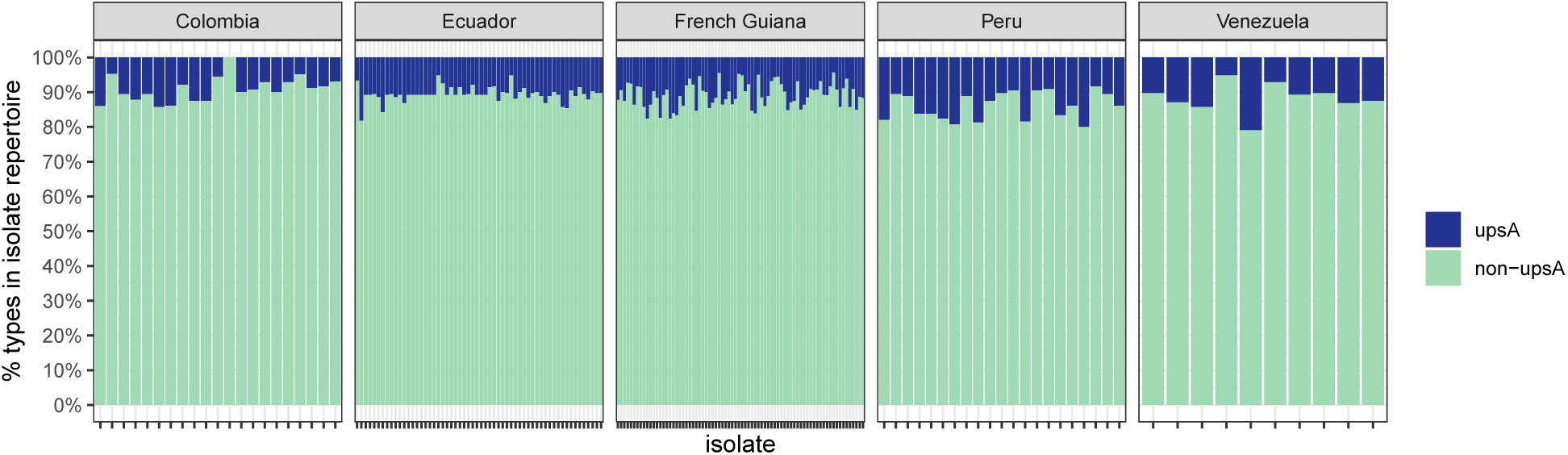
The proportion of upsA and non-upsA types in each isolate repertoire stratified by country.

**Figure 5:**
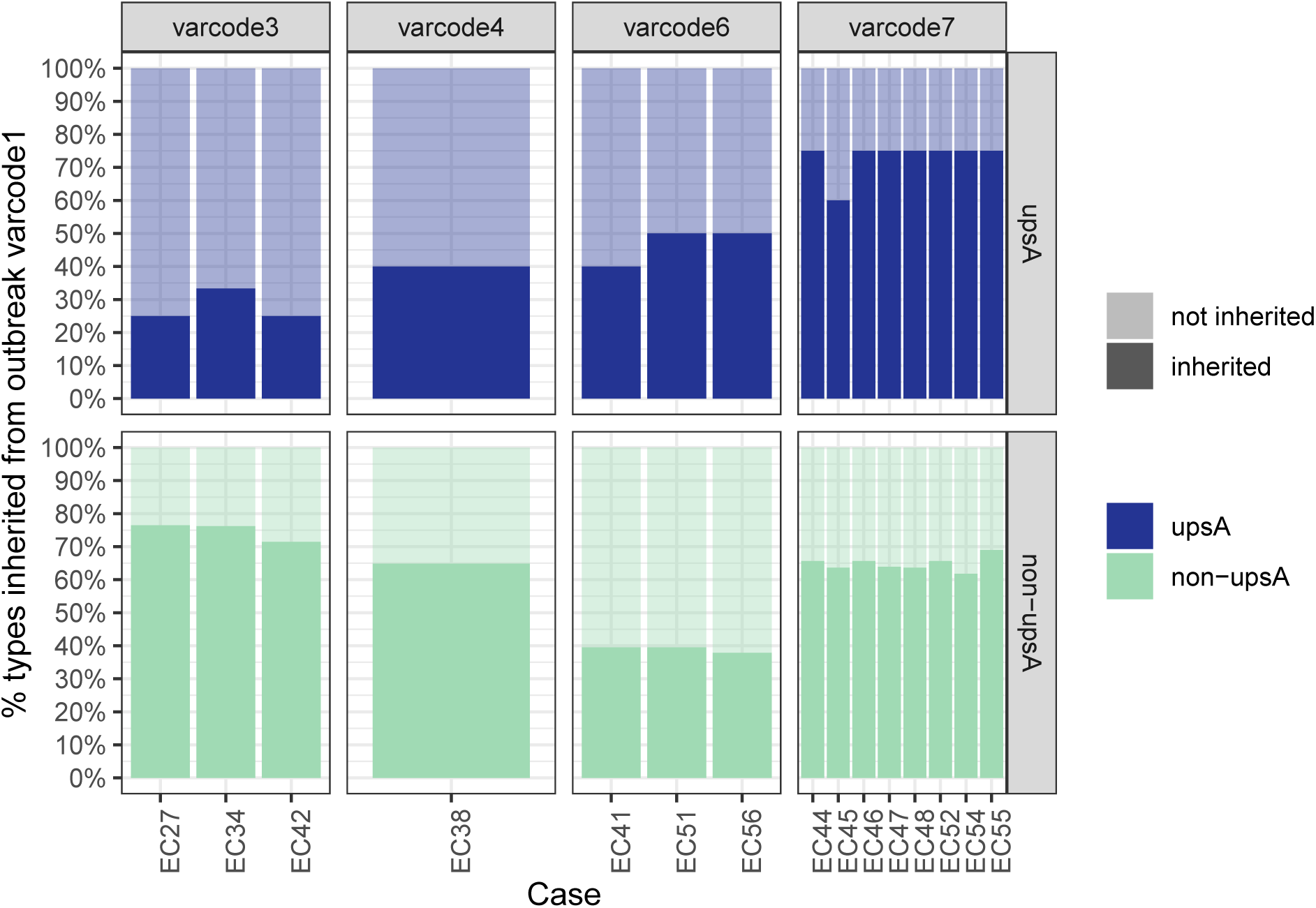
The proportion of the outbreak types that were inherited in the recombinant parasite varcodes is indicated in the darker shades of blue or green. The proportion of the outbreak types that were inherited in the recombinant parasite varcodes is indicated in the darker shades of blue or green and the lighter shades correspond to those types that were not inherited from the outbreak clone but from the other parent.

